# Childhood deprivation and Health related quality of life and associated factors among Pediatric cancer patients at National Hospital, Tanzania

**DOI:** 10.1101/2022.10.26.22281579

**Authors:** Mwanaheri Chubi, Stephen Kibusi, Lulu Chirande, Shakiru Juma

## Abstract

**Introduction:** Pediatrics cancer is one of the most important life-threatening, non-communicable diseases worldwide. However, pediatric cancer patients suffer from physical disabilities associated with cancer treatment. However, there is unclear information about level of health-related quality of life and associated factors. Aimed of this study was to assess level of Health-Related Quality of Life among pediatric cancer patients.

**Methods:** A hospital-based Analytical cross-sectional study design with a quantitative approach was employed among 91 conveniently sampled pediatric cancer patients with their parents/caregivers. Self-administered questionnaires were used to collect data. Data were analyzed using SPSS computer software program version 25. One-way ANOVA and linear regression were used to quantify and establish an association between Childhood Deprivation, Health-Related Quality of Life, and associated factors among Pediatric Cancer Patients at 95% CI, and a 5% level of significance.

**Results:** Results, the response rate was 92.3% (n = 84). The most prominent kind of pediatric cancer was Acute Lymphoblastic Leukemia 30 (35.7%) and Hodgkin lymphoma 11(13%). The mean age of pediatric cancer patients was 10.26±3.90 years while 59 (70.2%) were male. In linear regression, Childhood Deprivation (β=-1.640; P-value< 0.05) on PedsQL™ 4.0 and (β= -2.175; P-value< 0.01) on PedsQL™ 3.0. Findings imply that decreases the level of Childhood Deprivation, the chance of improving Quality of life increases.

**Conclusion:** The magnitude of acute lymphoblastic leukemia is higher among pediatric cancer patients. There was a significant link between pediatric cancers and childhood deprivation and thus, compromised quality of life. Innovative pediatric cancer care policies, guidelines, and or strategies may need to be advocated to address the problem accordingly.

## Introduction

Pediatrics cancer is one of the most important life-threatening non-communicable diseases worldwide. It has been predicted that more than 1,000 children will be diagnosed with cancer every day and 400,000 children will be diagnosed with cancer every year, most living in low- and middle-income countries [1]. Pediatric cancer patients are individuals who have been diagnosed with cancer from 0 to 18 years [2] with a survival rate of more than 80% in high-income countries and 5 to 25% in low and middle-income countries respectively [3].

However, little is documented about pediatric cancer patients in sub-Saharan Africa as there is no population database of pediatric cancers in Africa, making it difficult to know the true prevalence of pediatric cancers [4]. estimated a mortality rate of 4.6/100,000, from pediatric cancer with an incidence rate of 0.5 as high, with the mortality rate being twice as high for low- and middle-income countries where they are disadvantaged in terms of education, health, and standard of living [5,6].

Although evidence shows that the survival rate has significantly increased following the advancement of pediatric cancer therapies such as multiple chemotherapies, surgery, radiotherapy, supportive care, and bone marrow transplantation [7]. Furthermore, studies found that childhood cancer has the best 5 years relative survival rate after diagnosis and a high rate in children with retinoblastoma and ALL tumor types had more than 50% survival in Nairobi compared to Zimbabwe and Uganda, (8), and less than 18 months survival rate to Burkitt lymphoma in Tanzania [9]. Nevertheless, the prompt number of a male has been observed in many countries compared to female pediatric cancer patients most of them spend much time receiving cancer treatment approximately six months to 3 years [10], after diagnosis.

In Tanzania, 18.8% of pediatric cancer patients were diagnosed in 2019, with an incidence of 1.42/100,000 whereby Wilms tumor was 17.2%, retinoblastoma 16.8%, and acute lymphoblastic leukemia was 15.9%, furthermore, Burkett lymphoma and Wilm’s tumor has been presented mostly in lake zone region [11]. The World Health Organization has launched the Global Initiative for Childhood Cancer (GICC) to maximize the survival rate by up to 60% by 2030, worldwide [12].

Despite the substantial gains in recent years, due to major support rendered by developed countries and various international organizations, to support the government in the eradication of child poverty, evidence shows that human basic needs are deprived and significantly remain a major health problem among children [13]. However, there is limited published data about childhood deprivation in pediatric cancer patients worldwide, few studies reveal that in pediatric cancer patients living in the most deprived areas at diagnosis [14], and associated with poor quality of life. Furthermore, evidence shows that most pediatric patients who were diagnosed with cancer disease are hard to reach cancer treatment centers early as they should travel a long distance for care [11].

In Tanzania, about 88.9% of children suffer from more than three deprivations and half of them suffer from more than five deprivation including insufficient nutrition, health, protection, education, information, sanitation, water, and housing [15]. Furthermore, studies found that children living in poverty face many deprivations to their human rights, survival, health and nutrition, education, and participation [16]. In addition, a study done in Malaysia revealed that children living in poor (housing, and living conditions) and scarcity health care delivery are significantly vulnerable to physical, social, school, and emotional problems [17] which is among of the factors associated to lower mean scores of health related quality of life of pediateic cancer patients.

Health-Related Quality of life is defined as perceived physical, emotional, mental, and social well-being that is affected by a disease condition and its treatment [18]. WHO, Global Initiative Childhood Cancer on its strategies approach to cure all, based on improving health-related quality of life by maximizing the abilities of pediatric cancer patients to achieve individual goals while reaching productive fellows of society [4]. Furthermore, it is expected that the pediatric cancer patient should be free from the major physical and psychological challenges which will improve health related quality of life by alleviating those effects of cancer treatments such as learning difficulties, growth, and cognitive problems [19], by maximizing early interventions

Studies found that age at diagnosis, sex, place of residence, type of cancer disease, medical treatment, [20,21], level of education, marital status of the parents/caregiver, employment, poor housing and living standards [22], impact the perceived Health Related Quality of Life of pediatric cancer patients and their parents/caregivers in both generic core and and specific cancer module. Where Pediatrics Quality of Life Inventory (PedsQLTM) Generic core 4.0 measures all pediatrics with various chronic diseases and Cancer module 3.0 is the specific for pediatric cancer patients and measure the severity of the disease [18,23,24].

So far, some study shows there is a large disparity in quality of life between high and low-income countries. Evidence found that there is significant variation in HRQOL perceived by pediatric cancer patients and their parents on PedsQL scores between countries, whereby the US reported high scores for physical, emotional, and school functioning [25]. Despite the increased incidence of pediatric cancer, childhood deprivation and Health-Related Quality of Life (HRQoL) among pediatrics diagnosed with cancer are not known in Tanzania. This implies that there is a need to know the status of childhood deprivation and Health-related quality of life and associated factors to pediatric cancer patients in Tanzania. The importance of this study is to highlight Sustainable Development Goal number 3 (SDG3), sub-goal 3.1 (Health and well-being), which is to ensure healthy living and promote well-being for all people of all ages.

Tanzania is one of the developing countries with an increasing number of pediatric cancer patients in a situation MOHCDGEC has launched several initiatives to address the burden. These initiatives include modern and accurate diagnostic technologies and multimodal therapy including chemotherapy, radiotherapy, surgery and rehabilitation services to improve the quality of life of all pediatric cancer patients to ensure all requirements are easily accessed. Currently, 9 Hospitals in Tanzania provide cancer therapy including Muhimbili National Hospital which provides all modalities that are subject to increases in the survival rate of pediatric cancer patients from 0 to 5 years, where about 50% of all children diagnosed with cancer in 2019 survive, [26,27].

However, challenges related to diagnosis and treatment are critical, pediatric cancer patients experience stressful physical and emotional suffering that could greatly affect their well-being including lack of self-esteem, physical incapacity, and poor educational progress [28]. Furthermore, they are subjected to fear, anxiety, and depression and also weakening of immunity [28,29], related to pain, nausea and vomiting, fatigue, weakness, loss of limb function, attention deficit problems and loss of hair while on the cancer treatment period [28]. Hence if the problem is left unattended, increases long-term effects and mortality. Furthermore, most of the previous studies have been conducted to establish the prevalence, incidence, needs, and concerns of childhood cancer [3,9,11,29].

They helped to expose early, late and long-term effects of cancer treatment, risk of secondary malignancies, to risk of recurrence and also they confront many issues during the treatment period [30], which include childhood deprivation and social problems. Furthermore, their experience has been associated with an increased risk of detrimental psychosocial effects impacting emotional health, socialization, educational and vocational achievement, and health care access [31].

Many questions remain unanswered regarding interrelationship issues related to Deprivation and Quality of Life in this unique group. Therefore, this study aims to assess Childhood Deprivation and Health-Related Quality of Life among pediatric cancer patients, which could help the health profession to initiate comprehensive and consistent early intervention that could improve Quality of Life.

## Methods and Materials

### Research design

It is a hospital-based, analytical cross-sectional study design with a quantitative research approach that was held to find the association between variables including childhood deprivation and HRQoL among pediatric cancer patients with their parents/caregivers

### Study setting

The study was conducted in the pediatric oncology unit at Muhimbili National Hospital, which is a National referral hospital located at Ilala, Dar es Salaam Tanzania (Eastern Indian Ocean coast of Africa), with 1,500 beds capacity, attending 1,000 to 1,200 outpatients per week and admitting 1,000 to 1,200 inpatients per week. The actual data was collected at the pediatric oncology unit whereby pediatric cancer patients were found from all over the country ranging from 0 to 18 years old. It is a hub of pediatric oncology in Tanzania, with the unit attending 300 to 350 new cancer children each year [26].

Furthermore, approximately more than 150 pediatric cancer patients attend follow-up visits at pediatric oncology units per month, this includes all from cancer Hostels and homes with a revisit plan either weekly or monthly depending on their progress and treatment schedule. The unit runs its clinic on 3 alternative days per week (Monday, Wednesday, and Friday). The study were conducted from 6^th^ may to 30^th^ June 2022.

### Study population

The primary subjects in this study were confirmed pediatric cancer patients aged 5 to 18 years who received cancer treatment at the oncology unit (clinic) after being reviewed by an oncologist and planned for treatment.

### Inclusion and exclussion criteria

All pediatric patients who confirmed they had cancer disease and were on cancer treatment at the oncology clinic were included in the study aged from 5 to 18 years, and understood and communicate verbal instructions.

### Sampling and Sampling procedure

The following formula adopted from Cochran (1977) was used to determine the minimum sample size (n) by mean that was required for this study. A convenient sampling technique was used to recruit pediatric cancer patients aged 5 to 18 years who already completed an induction course of cancer therapy (Tembe et al, 2021). The patient’s files were used to find the diagnosis, phase, and types of cancer treatment of the child before recruitment. As shown in Fig 1.

**Fig 1:**
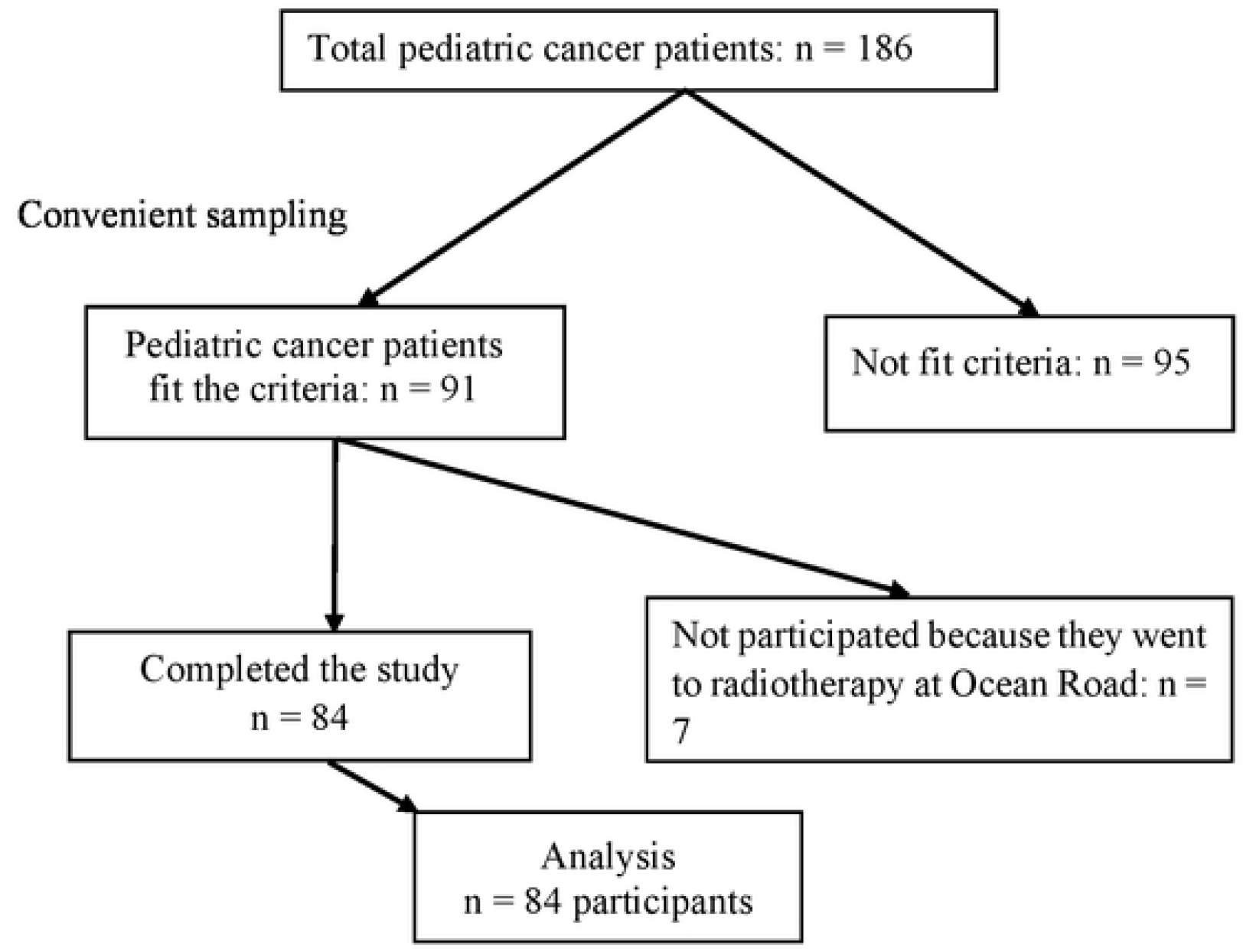
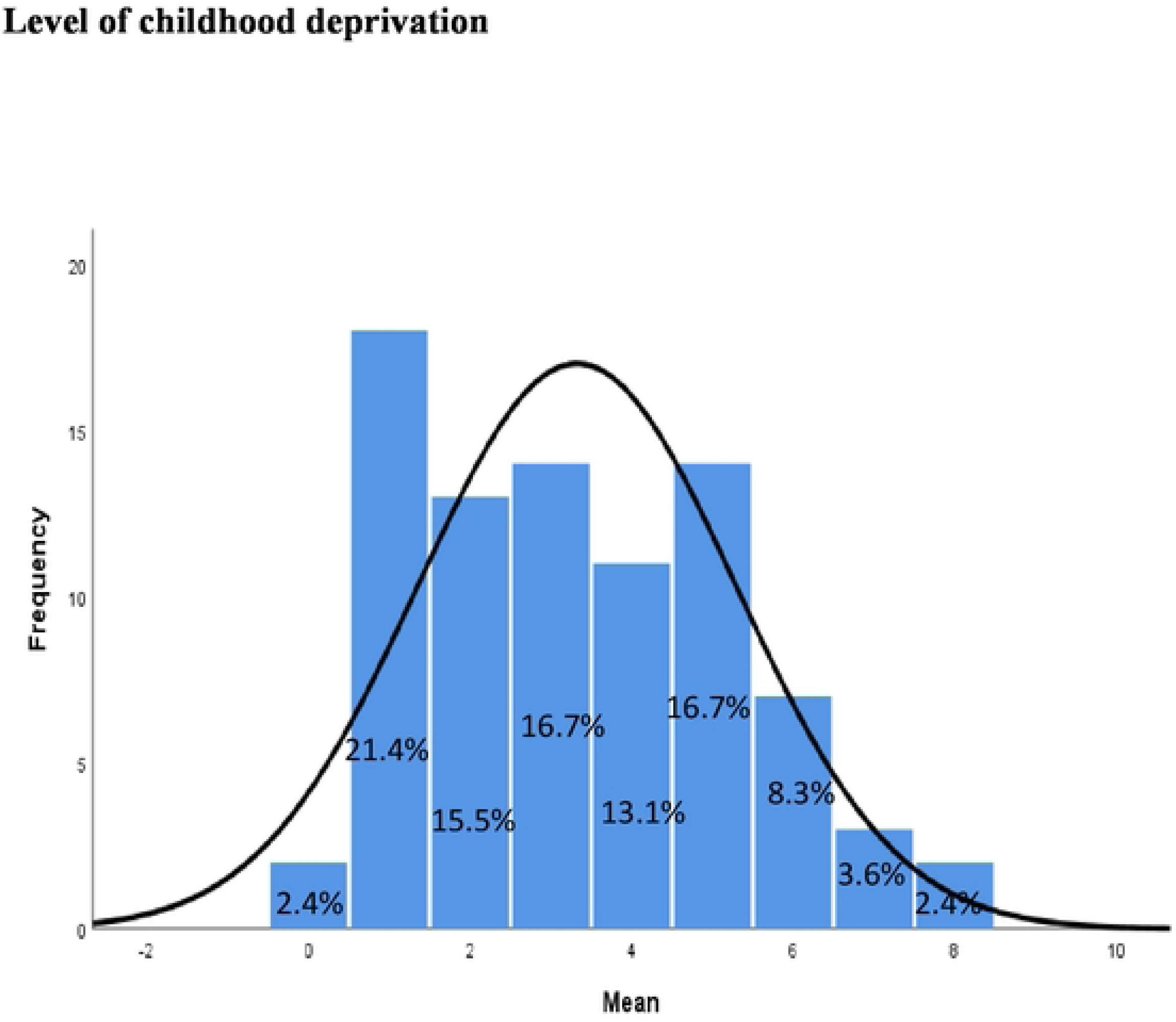
Flow diagram representing sampling procedure and sample size. Histogram showing the level of childhood deprivation by mean score among pediatric cancer patients

### Variables

#### Outcome variables

The two dependent variables were assessed including Health-Related Quality of Life and Childhood Deprivation among Pediatric cancer patients.

#### Predictors

The variables show the association on effect to dependent variable including social demographic data of Pediatric cancer patients and their Parents/caregivers and the disease profile

#### Data source

The study used and modefied the Social demographic characteristics from Tanzania maralia indicator survey of 2017 [32] and disease status from previous source [24] [18,24,34] and Multidimensional Poverty Index(MPI) was developed by Sabina Alkire and Foster in 2007, by using demographic and health survey data from USAID and from UNICEF (Multiple indicator cluster surveys (MICS) [16] and Pediatrics quality of Life inventory (PedsQL™ 4.0 Generic core scale and PedsQLTM 3.0 Cancer module) from American Pediatric Cancer Association [35]

### Measures and intrumentation

#### Social demographic characteristics

the data were collected using designed structured questionnaire, including pediatric cancer patients characteristic: Age, sex, level of education, place of residence and disease status. Parents/caregivers information including age, sex, level of education, employment status, marital status and commenced caring the child

#### Childhood deprivation

defined as lack of essential needs such as nutrition, water and sanitation facilities, shelter, access to basic health care services, education, participation, and protection leaving children not capable to enjoy the right to reach their full potentials as members of the community [32]. In this study, Global Multidimensional Poverty Index 2014 (MPI) scale used to measure poverty for catching multiple childhood deprivation including education, health and standard of living. Lack of essential needs among pediatric cancer patients resulting on unable to fulfill and participate various activities as a member of community.

The Global Multidimensional Poverty Index2014 (MPI) scale was used to examine each person’s deprivations across 10 with indicators in three equally weighted dimensions such health, education and standard of living (Nutrition, Child mortality, Cooking fuel, Sanitation, Drinking water, Electricity, Housing, Assets, Years of schooling and school attendance) Whereas each indicator is equally evaluated within its dimension, health and education indicators are weighted 1/6 each and the standard of living indicators are weighted 1/18 each. However, the normal MPI ranges from 0 to 1 (the domain was weight by using scale of 0 and 1) where 0 if the pediatric cancer patient fulfils the wealth criteria and 1 when does not fulfill wealth criteria. However, the standard of living involved 2 subscales one on item 6 and second on item 7 making a total of 12 questions.

#### Health Related Quality of life

defined as perceived physical, emotional, mental and social well-being is affected by a disease condition and its treatment, [18]. The Pediatric Quality of Life Inventory Version 4.0. (PedsQL™ 4.0) Generic core scale, used to assess the Quality of life of pediatric patients with cancer diseases. The scale comprises with 23-item with 4 components such as Physical Functioning score (8 items), Emotional Functioning score (5 items), Social Functioning score (5 items), and School Functioning score (5 items). The scales contained in both child report design include 5– 7 (young child), 8–12 (child), and 13–18 (adolescent).

The child report form for children completed as self-administered interview questions. (24,37). Items are scored on a 5-point response scale where by (0 = never a problem, 1 = almost never a problem, 2 = sometimes a problem, 3 = often a problem, 4 = almost always a problem). Whereas the specific domain and entire score is calculated from the equivalent questions, ranging from 0 to 100 (0=100, 1=75, 2=50, 3=25, 4=0). Furthermore, for ages 5-7 years’ items are scored on a 3-point response scale (0 = not at all, 2 = sometimes a problem, 4 = a lot of problems).

The PedsQL™ 3.0 cancer module will be used to obtain the subject’s information regarding HRQL. The module is a multidimensional instrument having eight domains which consist of 27 items that are covered by five scales: 1 (pain and hurt-two items),2 nausea (5 items), 3 procedural anxiety (3 items), 4 treatment anxiety (3 items), 5 worries (3 items), 6 cognitive problems (5 items), 7 perceived physical appearance (3 items) and 8 communication (3 items). Items are scored on a 5-point Likert scale whereby (0 = never a problem, 1 = almost never a problem, 2 = sometimes a problem, 3 = often a problem, 4 = almost always a problem for the children aged 8 to 18 self-report and parent proxy report. children aged 5 to 7 years have a 3-point Likert scale, where 0=not at all, 2= sometimes and 4 =a lot. The data was collected by using a visual aid (happy face, neutral face, and sad face) for ages 5-7 years, as a tool direction. Whereas the specific domain and entire score are calculated the same as in PedsQL™ 4.0 [35].

However, both Likert was transformed and shared with all age groups to 3 Likert scales 0=never a problem, 2 sometimes, and 4 almost always a problem where (0=100, 2=50, and 4=0). Furthermore, scores ranged from 27 to 135 for PedsQL 3.0 cancer module and 23 to 115 for PedsQL 4.0 Generic scale. Therefore, for this study, those who scored higher means have a better Quality of Life (above 69.7), and below are considered a poor quality of life [35,38], since there is no median severity of the disease

### Reliability of the study

On MPI, the Exploratory factor analysis was performed for reduction and determine weights of each item set at 0.3 as recommended by previous studies [37,38]. The correlation coefficient was set at a cut-off point of ≥0.30, whereas, the Kaiser-MeyerOklin (KMO) value was set at 50% with a 5% level of significance as shown in table 3.1. 11 items were subjected to an initial solution of which two items 1 and 4 were extracted as they weighed <0.3. The second exploratory factor analysis involved 9 items that weighed >0.3 and were for further analysis. The total variance was explained by factor analysis 3(initial Eugen value= 13.317 and the cumulative percent=63.646)

The descriptive analysis was done which was subjected to establish mean, standard deviation, percent, variance, range, and normality statistics including skewness and kurtosis. However, the data for childhood deprivation were approximately normally distributed, whereby Kolmogorov statistical value was 0.112(DF-84), P-value< 0.05 with skewness of 1.407, and kurtosis 1.381. Therefore, the non-parametric measurement was obtained and the mean was used as a central tendency to define the endpoint of childhood deprivation scores above the mean were treated as deprived or otherwise not deprived. As shown in table 3.1 mean score and standard of error for the cut point of level of childhood deprivation among pediatric cancer patients was 3.32(0.215), with a minimum of 0 and a maximum of 8.

The Pediatric Quality of life, the actual Cronbach alpha for the PedsQL™ 4.0 Generic core scale was 0-0.7 and the PedsQL™ cancer module inventory were ranging from 0-0.7. The tool was tested before conducting the study to assess it consistently, by providing a -pre-testing questionnaire in a geographical location outside of the study area, which has been sampled in the study. 16 (10%) pediatric patients with sickle cell disease were interviewed by using MPI, PedsQL™ 4.0 and PedsQL™ 3.0, was tested by using the Cronbach alpha values, which measured the internal consistency. The place of residence was used to test the tools. Both Health-related Qualities of life were excellent at Cronbach alpha 0.7. However, the MPI was excellent with Cronbach alpha ranging from 0.3-0.72 points in factor analysis after increases of ilitaration to 75% in the first round, because the sample size was below 300 participants.

### Sample size

The following formula adopted from Cochran 1977 was used to determine the minimum sample size (n) that was required for this study

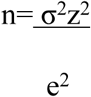

Whereas

n = Minimum sample size

σ = standard deviation (13.965 for the children aged≥ 5 years (18)

z= significant level 1.96

*e* = acceptable sampling error (3%)

Therefore

N = (13.965×1.96/3)^2^

N was 83 with an attrition rate of 10% of no response, therefore the final calculation was 91.

### Analytical strategy

Obtained data was managed by manually cross checked for completeness and accuracy to detect any missing information and was entered to SPSS version 25 so as to clean the data by running frequency, and to view if it is skewed or not skewed to detect any extreme value and opt the type of data analysis model. Descriptive statistics were performed to establish social demographic characteristics of pediatric cancer patients and their parents/caregivers and also were used to find the level of childhood deprivation and health related quality of life.

Cross tabulation of frequence and percentage of socio demographic charactreistics and disease profile were performed. One-way ANOVA and independent t-test two tailed was used to find the diference between variables of sociademographic characteristics and outcome variables. Linear regression was used to investigate the association between outcome variable and variable of interest including socio demographic characteristics and disease profile.

### Ethics approval and consent to participate

Permission to conduct the study was brought from the University of Dodoma through the Institution Research Review Committee (IRRC), and the Executive director of MNH to allow the study to be conducted at the oncology unit. The informed consent form was obtained from parents/caregivers and assent was obtained from pediatric cancer patients by the Principle Investigator before being interviewed. No names of participants was appearing on the questionnaire, only numbers were coded to ensure confidentiality. The data collection procedure was conducted in an unoccupied separate room to ensure privacy. The participants had the right to withdraw from the study. However, during the study, no challenge or any change arises regarding Health Related or childhood deprivation among pediatric cancer patients. Data was handled and managed securely by using a folder with a key in the IRRC computer and not shared with other users for any purpose out of the intention of this study.

## Results

### Socio-demographic information of the pediatric cancer patients

Pediatric cancer patients and their parents/carers qualified and participated in the study with a 92.3% (n=84) response rate of 91. As shown in Table 1, the mean age of 84 pediatric cancer patients was 10,263,90 with a minimum of 5 and a maximum of 18 years The most prominent age group ranged between 8 and 12 years 40.5% (n=34), as most of them were male 70.2% (n=59) compared to females 29.8% (n=25). Most of pediatric cancer patients living in urban 38.1% (n=32), Rural 36.9% (n=31) and Peri-Urban 25% (n=21). Only 47.6% (n=40) of them attended school during the study compared to 52.4% (n=44) of them are not going to school, while about 63.1% (n=53) had primary school level.

**Table 1.**
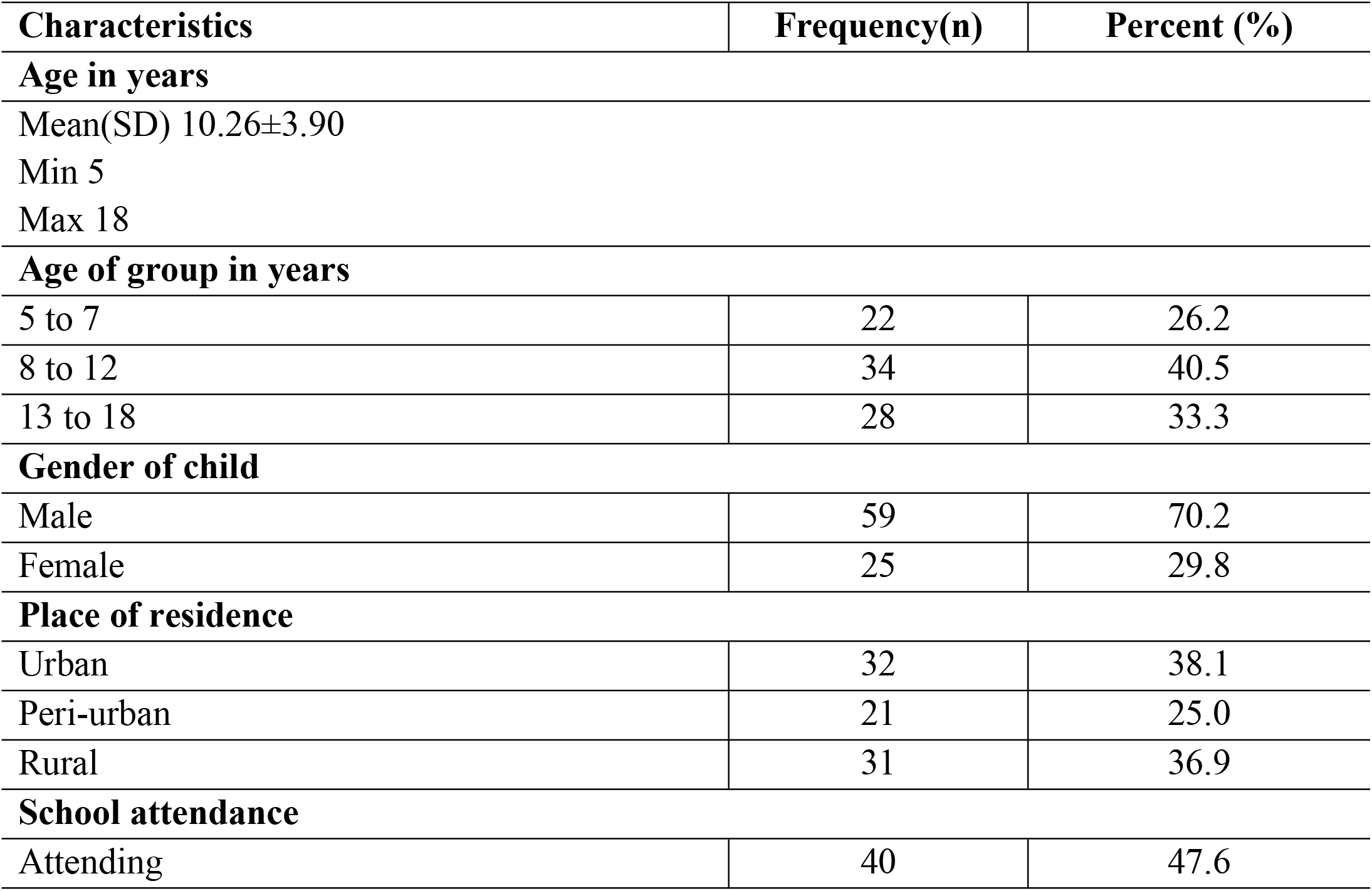
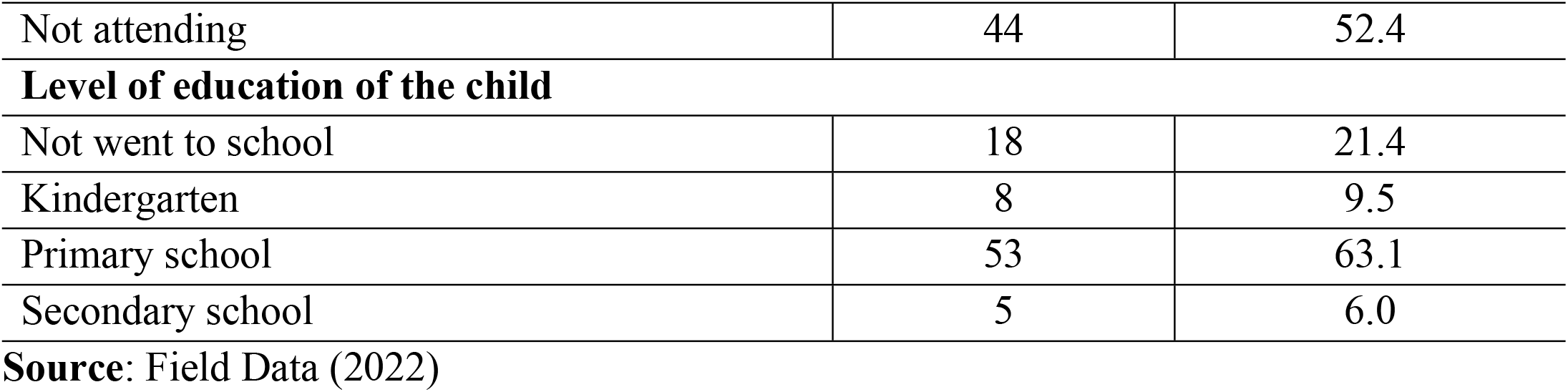
Socio-demographic information of the pediatric cancer patients (n=84)

### Socio-demographic characteristics of parents/caregivers

Table 2 shows that the mean age of 84 qualified parents/carers who participated in the study was 38.6210.87 years, with a minimum of 18 and a maximum of 68 years. The most conspicuous 58.3% (n=49) of parents/caregivers aged 20-39, 52% (n=44) were female and 47.6% (n=40) were male, 41.7% (n =35) were mothers. Also whom 71.6% (n=61) were married, 58.3% (n=49) had elementary school, 85.7% (n= 72) were informally employed and about 78.6% (n=66) started caring for the child before the illness. The results may indicate that the majority of caregivers are young and capable of caring for pediatric cancer patients and influencing the patient’s good health.

**Table 2.**
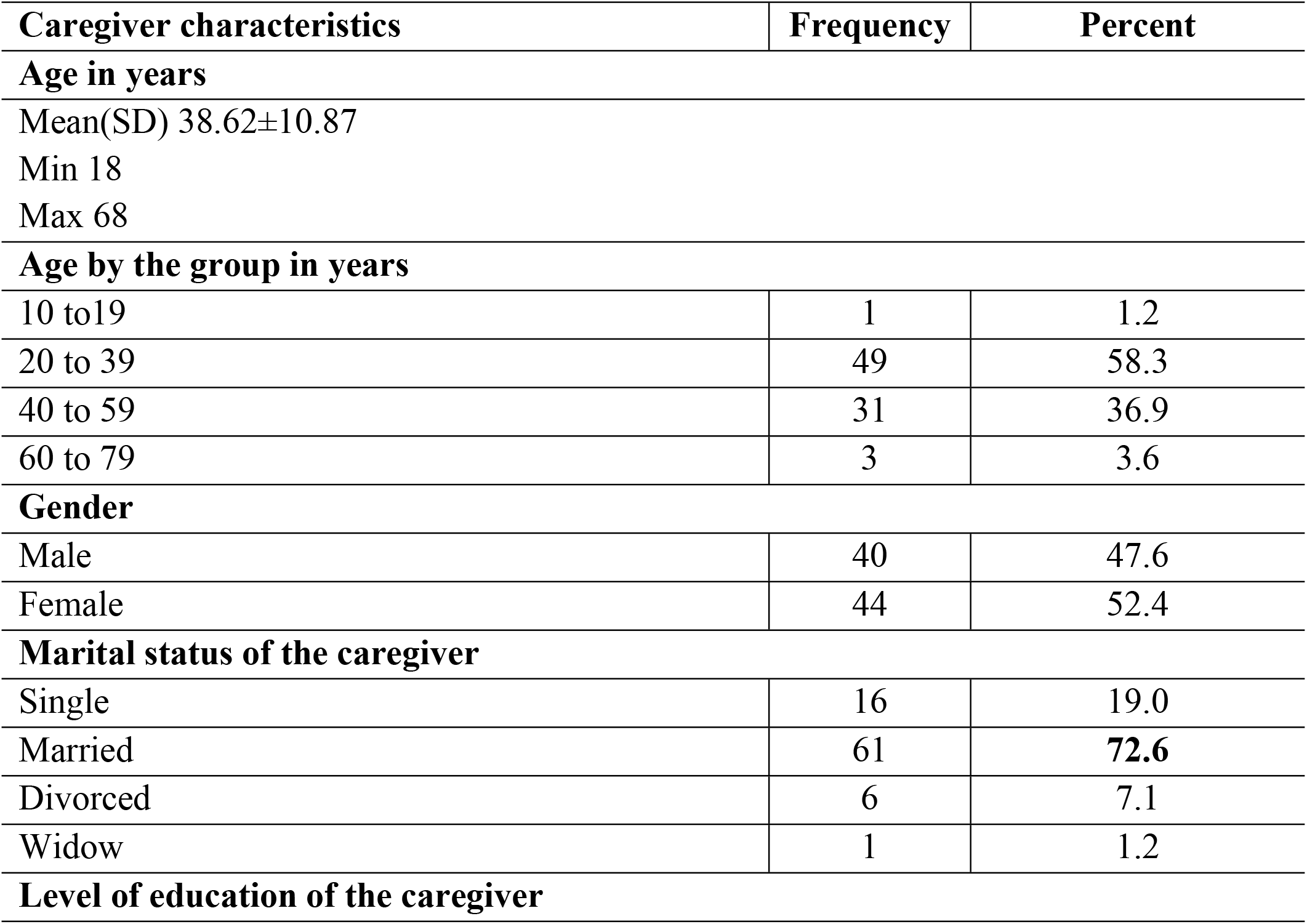
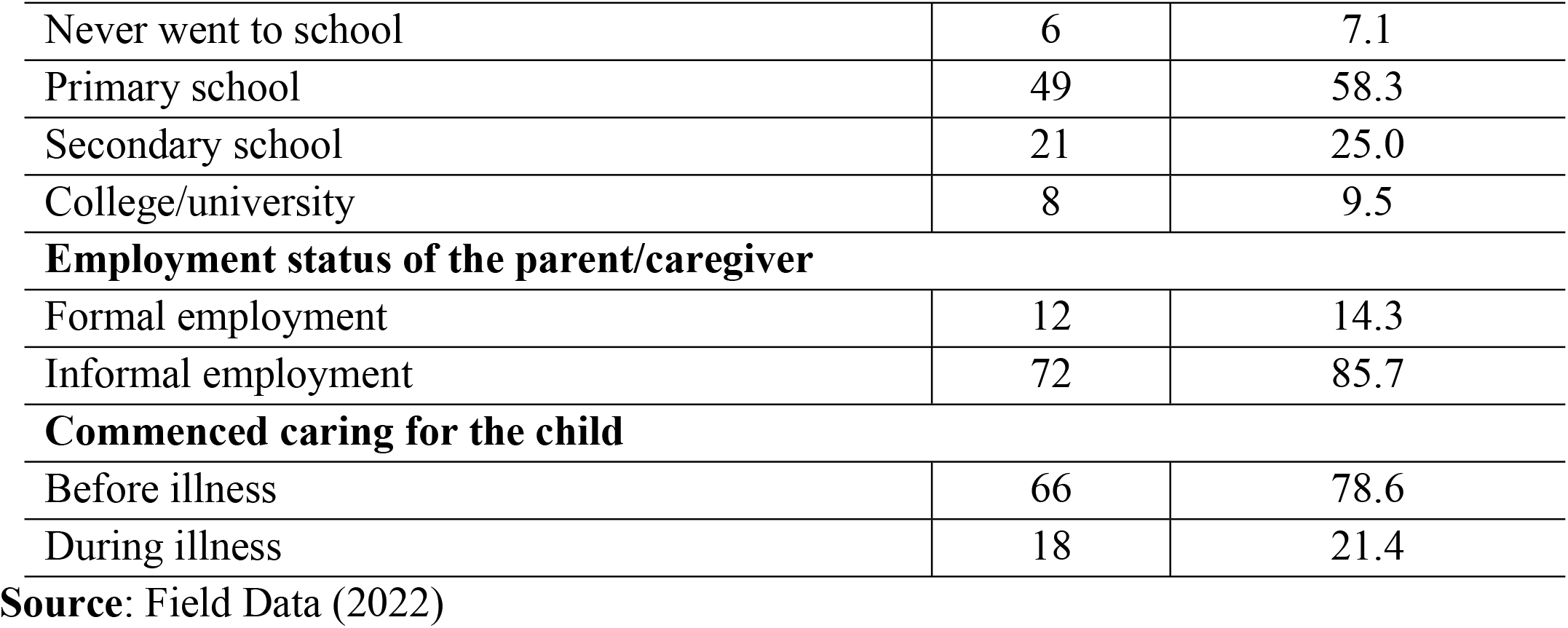
Socio-demographic characteristics of Parents/caregivers (n=84)

### Disease profile of pediatric cancer patients

As shown in Table 3, the results show that the most prominent pediatric cancer was 35.7% (n=30) acute lymphoblastic leukemia, followed by 13.7% (n=11) Hodgkin’s lymphoma, and the last one for rhabdomyosarcoma, one for primitive neuroectodermal tumor, one for endometrial carcinoma and one for esophageal carcinoma. Tumor while most of them confirmed cancer and started cancer treatment in MNH 89.3% (n=75). The median year of diagnosis was 2019,772.47 with a minimum of 2009 and a maximum of 2022, with the majority of pediatric cancer patients 63.1% (n=53) diagnosing cancer in 2021 and early 2022.

**Table 3.** Disease profile of pediatric cancer patients (n = 84)

| Characteristics | Frequency | Percent |
| --- | --- | --- |
| **Year of diagnosis** |  |  |
| Mean(SD); 2019.77 ±2.47 |  |  |
| Min 2009 |  |  |
| Max 2022 |  |  |
| **Year of cancer disease diagnosed** |  |  |
| 2009 | 1 | 1.2 |
| 2010-2015 | 2 | 2.4 |
| 2016-2020 | 28 | 33.3 |
| 2021-2022 | 53 | 63.1 |
| **Time since diagnosis in years** |  |  |
| Mean(SD); 1.89 ±0.99 |  |  |
| Min 1 |  |  |
| Max 6 |  |  |
| **Time since diagnosis by group(years)** |  |  |
| <1 year | **38** | **45.2** |
| 1 to 2 years | 25 | 29.8 |
| 3 to 4 years | 13 | 15.5 |
| ≥5 years | 8 | 9.5 |
| **No hospitals treated the child before diagnosis** |  |  |
| 1 | 9 | 10.7 |
| 2 | 24 | 28.6 |
| 3 | 21 | 25.0 |
| ≥4 | 30 | 35.7 |
| **Diagnosis** |  |  |
| Acute Lymphoblastic Leukemia | 30 | 35.7 |
| Hodgkins Lymphoma | 11 | 13.1 |
| Cellular carcinoma with Xeroderma Pigmentosum | 10 | 11.9 |
| Wilm's Tumor | 9 | 10.7 |
| Burkitt Lymphoma | 8 | 9.5 |
| Retinoblastoma | 5 | 6 |
| Acute Amyloid Lymphoblastic | 4 | 4.8 |
| Osteosarcoma | 3 | 3.6 |
| Rhabdomyosarcoma | 1 | 1.2 |
| Primitive Neuroectodema tumor | 1 | 1.2 |
| Endometrial Carcinoma | 1 | 1.2 |
| Oesophageal tumor | 1 | 1.2 |
| **Cancer medication started after diagnosis** |  |  |
| ≥ 2 weeks | 60 | 71.4 |
| < 2 weeks | 24 | 28.6 |
| **Treatment** |  |  |
| Chemotherapy | 66 | 78.6 |
| Combination (chemotherapy, radiotherapy, and surgery) | 7 | 8.3 |
| Combination (chemotherapy and radiotherapy) | 2 | 2.4 |
| Combination (chemotherapy and surgery) | 9 | 10.7 |
| **Hospitalization in prior 3 months** |  |  |
| 1 to 3 times | 46 | 54.8 |
| >3 times | 35 | 41.7 |
| No hospitalization | 3 | 3.6 |
| **Psychosocial counseling** |  |  |
| Received | 62 | 73.8 |
| Not received | 22 | 26.2 |
| **Activities done in the last 3 months** |  |  |
| School/studying | 37 | 44.0 |
| Playing/exercises | 23 | 27.4 |
| Hospitalized | 12 | 14.3 |
| Too ill | 9 | 10.7 |
| Hand capped/ cannot work | 3 | 3.6 |
| **Complications related to cancer treatment** |  |  |
| Stomach pain | 36 | 42.9 |
| Fever | 31 | 36.9 |
| Heart palpitation | 7 | 8.3 |
| Anemia | 3 | 3.6 |
| Combination; Fever, stomach pain, heart palpitation, loss of appetite, and body malaise | 2 | 2.4 |
| Lack of appetite and fatigue | 2 | 2.4 |
| Diarrhea | 1 | 1.2 |
| Pneumonia | 1 | 1.2 |
| None | 1 | 1.2 |

**Source:** Field Data (2022)

The mean time since diagnosis was 1,890.99 with a minimum of 1 and a maximum of 6 years in pediatric cancer patients, while most of them were 1 year 45.2% (n=38), 1 to 2 years 29.8% (n= 25), 3 to 4 years 15.5% (n = 13) and 5 years are only 9.5% (n = 8) since confirmed disease. About 35.7% (n=30) of them were treated in 4 hospitals before diagnosis, 76.2% (n=64) started cancer treatment 2 weeks after initial diagnosis, 78.6% (n=66) had received chemotherapy, 73.8% (n=66) had received chemotherapy, 73.8% (n=64) had received cancer treatment =62) psychological support. However, 54.8% (n=46) of pediatric cancer patients hospitalized 1 to 3 times before 3 months, 42.9% (n=36) had stomach pain, and 36.9% (n=31) had fever, while receiving cancer treatment. Only 44.0% (n=37) of pediatric cancer patients attended school/university in the last 3 months.

The results imply that the number of children diagnosed with cancer is currently still increasing, while the majority of them delayed the start of cancer treatment, as most of them confirmed cancer and started cancer treatment at the National Referral Hospital, where all modalities of cancer therapy are available. However, most pediatric cancer patients experience stomach pain as the main problem while receiving cancer therapy, especially chemotherapy.

### The proportion of the type of pediatric cancer by sex,age and hospitalization prior to 3 months

Descriptive analysis was performed to determine the proportion of cancer type by gender, age group and hospitalization prior to 3 months in pediatric cancer patients as shown in table 4-6. The findings show that males were most affected at 70.2% (n=59) compared to females. In addition, ALL was 45.8% (n=27) in males and 12.0% (n=3) and Hodgkin’s lymphoma 15.3% (n=9) in males and 8.0% (n=2) most prominent in female pediatric cancer patients. In addition, the least cancer type was affected only in female pediatric cancer patients, including 4.0% (n = 1) Primitive Neuro-Ectodermal tumors (PNET), 4.0% (n = 1) endometrial cancer, and 4.0% (n = 1) Esophageal tumor.

**Table 4.** The proportion of diagnosis by age group (years) of pediatric cancer patients n=84.

| Variable | ALL | Retino<br>brastobl<br>astoma | wilm's<br>tumor | hodgikins<br>lymphom<br>a | Burkitt<br>lympho<br>ma | Osteo<br>sarcoma | Rhabd<br>o<br>myosar<br>coma | Cellular<br>carcinoma<br>with<br>xeroderma<br>pigmentosa | AML | PNET | Endom<br>etrial<br>carcino<br>ma | oesophage<br>al tumor | Total |
| --- | --- | --- | --- | --- | --- | --- | --- | --- | --- | --- | --- | --- | --- |
| 5 to 7 | 5(22.7%) | 2(9.1%) | 5(22.7%) | 5(22.7%) | 2(9.1%) | 0(0.0%) | 0(0.0%) | 2(9.1%) | 1(4.5%) | 0(0.0%) | 0(0.0%) | 0(0.0%) | 22(26.2%) |
| 8 to 12 | 13(38.2%) | 1(2.9%) | 4(11.8%) | 4(11.8%) | 2(5.9%) | 0(0.0%) | 1(2.9%) | 5(14.7%) | 2(5.9%) | 1(2.9%) | 1(2.9%) | 0(0.0%) | 34(40.5%) |
| 13 to 18 | 12(42.9%) | 2(7.1%) | 0(0.0%) | 2(7.1%) | 4(14.3%) | 3(10.7%) | 0(0.0%) | 3(10.7%) | 1(3.6%) | 0(0.0%) | 0(0.0%) | 1(3.6%) | 28(33.3%) |
| Total | 30(35.7%) | 5(6.0%) | 9(10.7%) | 11(13.1%) | 8(9.5%) | 3(3.6%) | 1(1.2%) | 10(11.9%) | 4(4.8%) | 1(1.2%) | 1(1.2%) | 1(1.2%) | 84(100%) |

**Table 5.**
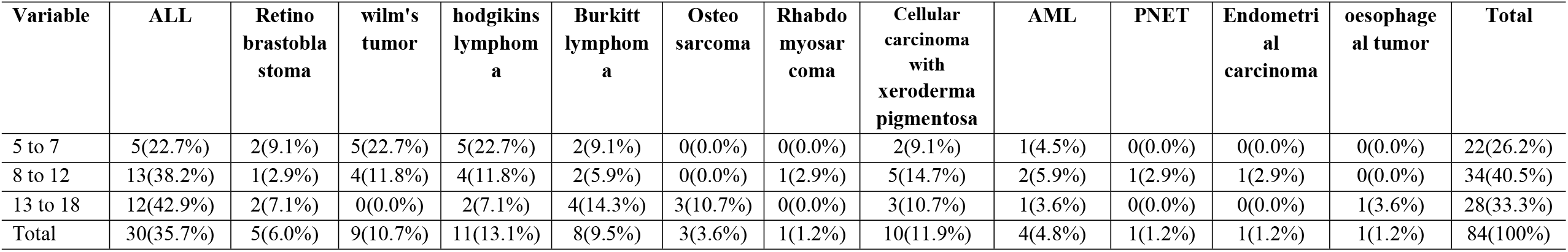
The proportion of diagnosis by age group (years) of pediatric cancer patients n=84.

| Variable | ALL | Retino<br>brastobla<br>stoma | wilm's<br>tumor | hodgikins<br>lymphom<br>a | Burkitt<br>lymphom<br>a | Osteo<br>sarcoma | Rhabdo<br>myosar<br>coma | Cellular<br>carcinoma<br>with<br>xeroderma<br>pigmentosa | AML | PNET | Endometri<br>al<br>carcinoma | oesophage<br>al tumor | Total |
| --- | --- | --- | --- | --- | --- | --- | --- | --- | --- | --- | --- | --- | --- |
| 5 to 7 | 5(22.7%) | 2(9.1%) | 5(22.7%) | 5(22.7%) | 2(9.1%) | 0(0.0%) | 0(0.0%) | 2(9.1%) | 1(4.5%) | 0(0.0%) | 0(0.0%) | 0(0.0%) | 22(26.2%) |
| 8 to 12 | 13(38.2%) | 1(2.9%) | 4(11.8%) | 4(11.8%) | 2(5.9%) | 0(0.0%) | 1(2.9%) | 5(14.7%) | 2(5.9%) | 1(2.9%) | 1(2.9%) | 0(0.0%) | 34(40.5%) |
| 13 to 18 | 12(42.9%) | 2(7.1%) | 0(0.0%) | 2(7.1%) | 4(14.3%) | 3(10.7%) | 0(0.0%) | 3(10.7%) | 1(3.6%) | 0(0.0%) | 0(0.0%) | 1(3.6%) | 28(33.3%) |
| Total | 30(35.7%) | 5(6.0%) | 9(10.7%) | 11(13.1%) | 8(9.5%) | 3(3.6%) | 1(1.2%) | 10(11.9%) | 4(4.8%) | 1(1.2%) | 1(1.2%) | 1(1.2%) | 84(100%) |

**Table 6.**
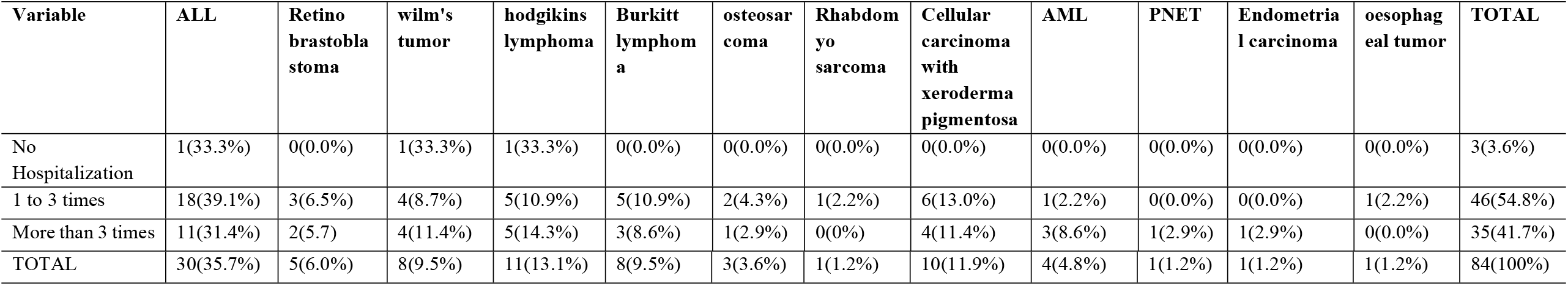
The proportion of diagnosis by hospitalization before 3 months of pediatric cancer patients n=84.

The proportion of cancers by age group. Results showed that pediatric cancer patients aged 8-12 and 13-18 years were most affected by ALL compared to other cancers, at 38.2% (n=13) and 42.9% (n=12), respectively. However, Hodgkin’s lymphoma and Wilms’ tumor affected most pediatric cancers at ages 5-7 and 8-12 years, with the same proportions of 22.7% (n=5) and 11.8% (n=4), respectively. Therefore, the most affected age group with pediatric cancer was between 8 and 12 years. In terms of hospitalization prior to 3 months, most pediatric cancer patients with ALL were admitted 1 to 3 times and more than 3 times, 39.1% (n=18) and 31.4% (n=11), respectively Hodgkin lymphoma and cellular carcinoma with xerodermatic pigmentosa as shown in Table.

### Level of childhood deprivation

A descriptive analysis was carried out, in which mean, frequency, and percentage were determined. As shown in the Fig 1, the data for the degree of childhood deprivation were approximately normally distributed, since all values above the mean were considered disadvantaged in pediatric cancer patients, since the cut-off value was 3.32. The results show that approximately 56.0% (n=47) of pediatric cancer patients were deprived.

### Mean scores of childhood deprivation by age

The overall mean values are below the limit in both age categories. However, the observed differences in standard of living where the mean is above the threshold are at ages 5-7 years (3.64±1.81) compared to ages 8-12 and 13-18 years (2.44±1.54) and (2.71±1.76) or as shown in Table 7. The results suggest that most young pediatric cancer patients have a lower standard of living compared to older patients.

**Table 7.** Mean scores of childhood deprivation by age n=84.

| Variable | 5-7 yrs. |  | 8-12 yrs. |  | 13-18 yrs. |  | P-value |
| --- | --- | --- | --- | --- | --- | --- | --- |
|  | Mean | SD | Mean | SD | Mean | SD |  |
| Total mean score | 1.39 | 0.91 | 0.97 | 0.79 | 1.04 | 0.86 | 0.331 |
| Education | 0.32 | 0.48 | 0.15 | 0.36 | 0.14 | 0.36 | 0.212 |
| Health | 0.23 | 0.43 | 0.32 | 0.48 | 0.29 | 0.46 | 0.746 |
| Standard of living | 3.64 | 1.81 | 2.44 | 1.54 | 2.71 | 1.76 | 0.036 |

### Bonferroni alpha Post hoc test of childhood deprivation among pediatric cancer patients for age n=84

Multiple comparison tests in pediatric cancer patients showed a difference in significance between those aged 5-7 years and 8-12 years for standard of living, with the p-value being <0.05. However, in other domains, the observed mean difference was not statistically significant, as shown in Table 8. The results reflect that young pediatric cancer patients may deprived on standard of living.

**Table 8.**
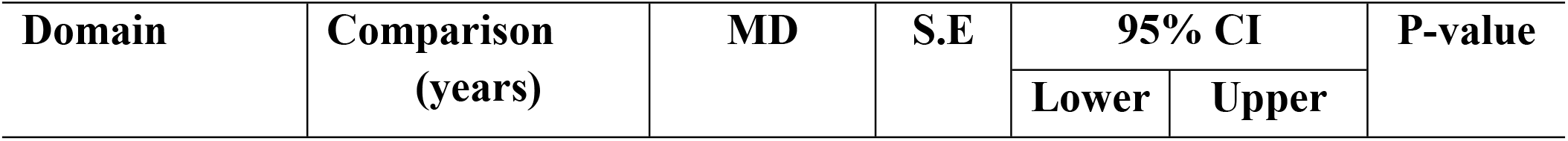
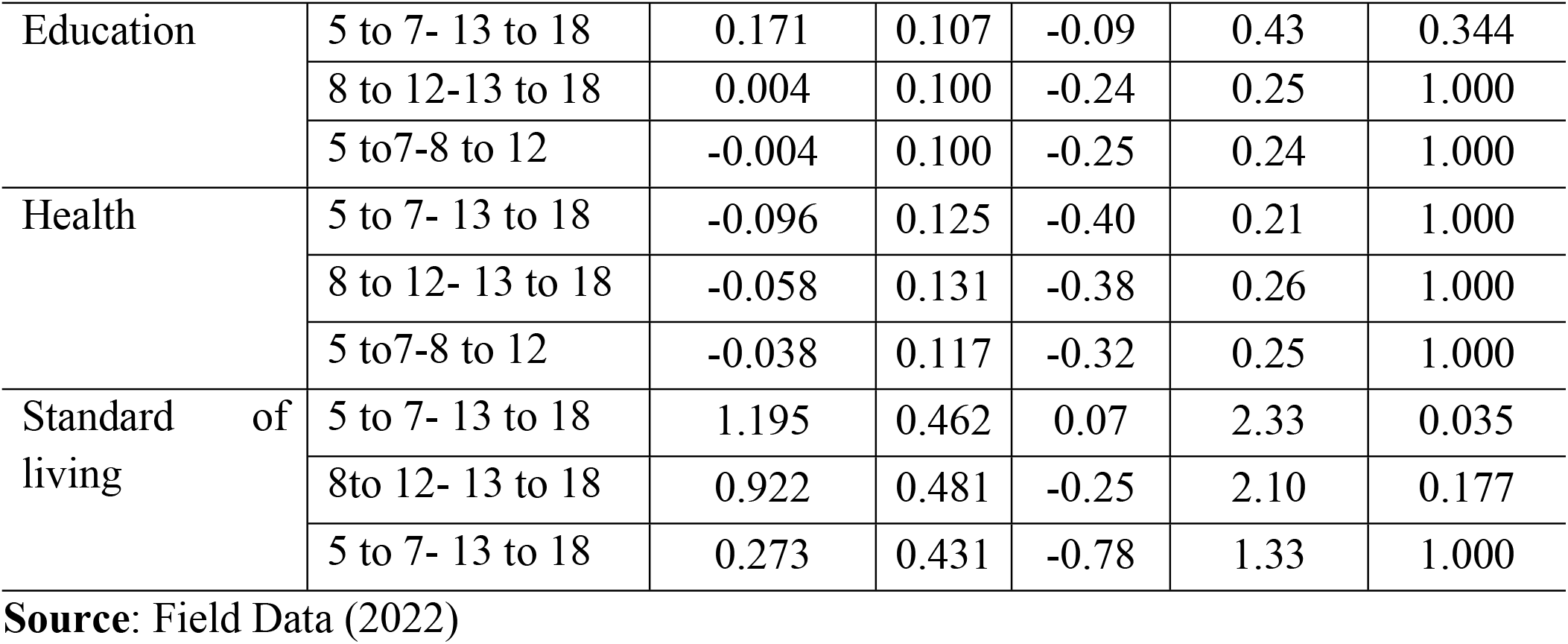
Bonferroni alpha Post hoc test of childhood deprivation among pediatric cancer patients for age n=84.

### Mean score of childhood deprivation by place of residence

As shown in Table 9, the mean total childhood deprivation scores by pediatric cancer patients’ city of residence. The results show that the overall mean is below the cut-off (3.32). However, mean living standard scores for those living in rural areas were higher (3.77±1.75; p-value <0.01) than those in urban and peri-urban areas. The finding implicates that living in rural areas could affect pediatric cancer patients, which resulted in poor quality of life.

**Table 9.** Mean scores of Childhood Deprivation by place of residence n=84.

| Variable | Urban |  | Peri-Urban |  | Rural |  | P-value |
| --- | --- | --- | --- | --- | --- | --- | --- |
|  | Mean | SD | Mean | SD | Mean | SD |  |
| Total mean score | 0.77 | 0.74 | 0.89 | 0.75 | 1.48 | 0.89 | 0.257 |
| Education | 0.16 | 0.37 | 0.24 | 0.44 | 0.19 | 0.40 | 0.765 |
| Health | 0.19 | 0.39 | 0.14 | 0.36 | 0.48 | 0.51 | 0.007 |
| Standard of living | 1.97 | 1.47 | 2.81 | 1.44 | 3.77 | 1.75 | 0.000 |

Source: Field Data (2022)

### Bonferroni alpha Post hoc test of childhood deprivation among pediatric cancer patients for place of residence

Multiple comparison tests in pediatric cancer patients showed a significant difference between those living in urban-rural and peri-urban-rural areas in terms of health functioning by the Bonferroni alpha post-hoc test with an adjusted p-value of <0.024 or 0.020. In terms of standard of living, the observed difference was between urban and rural. Therefore, the results suggest that pediatric cancer patients living in rural areas are disadvantaged in terms of health and standard of living. In addition, no difference was observed in education as shown in Table 10.

**Table 10.** Bonferroni alpha Post hoc test of childhood deprivation among pediatric cancer patients for place of residence n=84.

| Variable | Comparison | MD | S.E | 95% CI |  | P-value |
| --- | --- | --- | --- | --- | --- | --- |
|  |  |  |  | Lower | Upper |  |
| Education | Urban-peri urban | -0.082 | 0.112 | -0.36 | 0.19 | 1.000 |
|  | Urban-rural | -0.037 | 0.100 | -0.28 | 0.21 | 1.000 |
|  | Peri urban-rural | -0.045 | 0.113 | -0.32 | 0.23 | 1.000 |
| Health | Urban-peri urban | 0.045 | 0.122 | -0.25 | 0.34 | 1.000 |
|  | Urban-rural | -0.296 | 0.109 | -0.56 | -0.03 | 0.024 |
|  | Peri urban-rural | 0.341 | 0.122 | 0.04 | 0.64 | 0.020 |
| Standard of living | Urban-peri urban | -0.841 | 0.441 | -1.92 | 0.24 | 0.180 |
|  | Urban-rural | -1.805 | 0.396 | -2.77 | -0.84 | 0.000 |
|  | Peri urban-rural | 0.965 | 0.444 | -0.12 | 2.05 | 0.098 |

Source: Field Data (2022)

### Level of Health-Related Quality of Life and associated factors among pediatric cancer patients

The descriptive analysis was performed to determine the level of health-related quality of life (HRQoL), with the endpoint being the mean score from the tool such as the Pediatric Quality of Life Inventory, both the generic core scale (PedsQLTM 4.0) child report, while mean for child report (69.7) and parent proxy report (65.4). Therefore, the mean scores above the endpoint were treated with better quality of life and below the mean with poor quality of life

### Level of health-related quality of life among pediatric cancer patients measured by (PedsQL™ 4.0) Generic core scale

As shown in table 11, the results of PedsQL™ 4.0 show that the total mean scores were lower (67.79±21.63). This finding implies that pediatric cancer patients have a poor quality of life, as the overall mean scores were below the cut-off point (69.7).

**Table 11.**
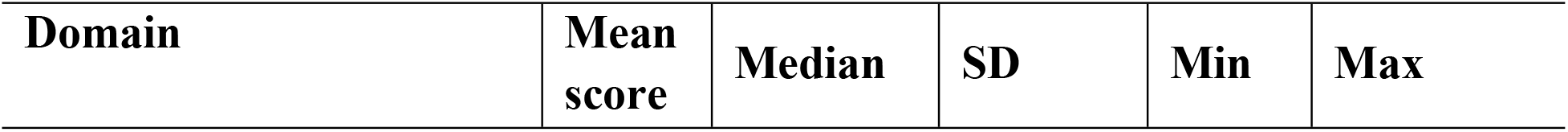
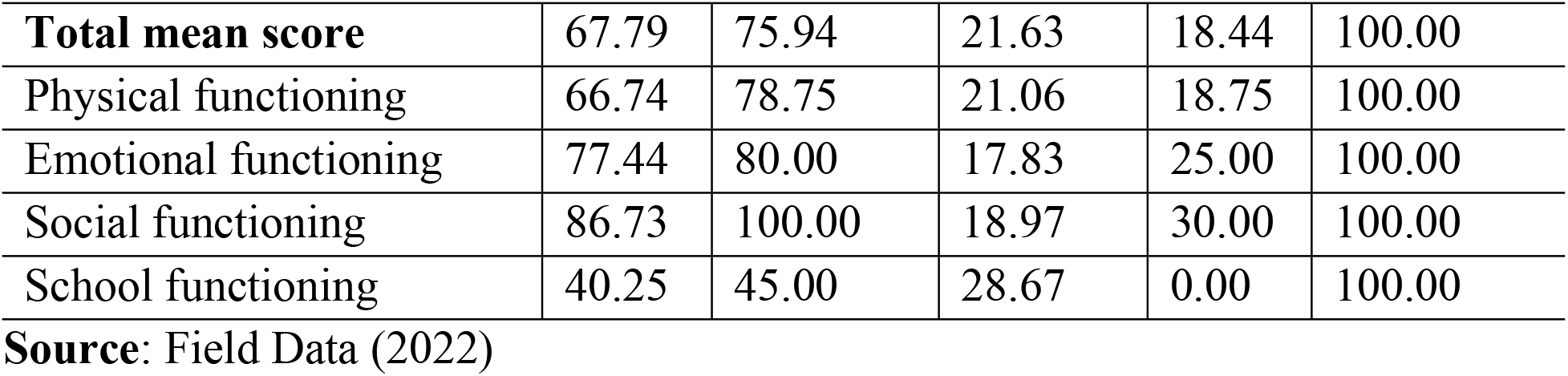
Total mean scores of the Level of Health-Related Quality Life measured by (PedsQL™ 4.0) Generic core scale (n = 84)

### Mean scores of Health-related quality of life measured by (PedsQL™ 4.0) Generic core scale by sex

Table 13 shows that the average total scores of males compared to females were below the cutoff point (69.7) in both the Child Report. However, only physical functioning was statistically significant (p-value 0.039). In addition, academic performance in both males and females was below the threshold but was not statistically significant. This finding suggests that the quality of life of female pediatric cancer patients improved and got better.

**Table 13.** Mean scores of Health-related quality of life measured by (PedsQL™ 4.0) Generic core scale by sex (n = 84)

| **Domain** | **Male** |  | **Female** |  | **P-value** |
| --- | --- | --- | --- | --- | --- |
|  | **Mean score** | **SD** | **Mean score** | **SD** |  |
| **Total mean score** | **65.46** | 21.85 | 72.08 | 21.35 | **0.313** |
| Physical functioning | **63.51** | 19.98 | 74.38 | 21.96 | **0.039** |
| Emotional functioning | 76.52 | 18.53 | 79.50 | 16.19 | 0.473 |
| Social functioning | 85.68 | 19.90 | 89.20 | 16.69 | 0.440 |
| School functioning | 38.14 | 27.82 | 45.24 | 30.57 | 0.302 |

**Source:** Field Data (2022)

### Mean scores of Health-related quality of life measured by (PedsQL™ 4.0) Generic core scale for age

Table 14 One-way ANOVA analysis was used to establish the relationship through mean results of the PedsQL 4.0 Generic Core Scale for Child report by age group. The results show that age groups (13-18 and 8-12 years) had a significant impact (M=70.00±15.32 and 73.33±10.31, respectively; p<0.01) on the health-related quality of life of pediatric cancer patients. However, the high mean was statistically significant for physical functioning at ages 8–12 and 13–18 years (M=71.59±19.09; 71.14±19.99; p-value 0.001) compared to the 5–7 year-olds who reported a low (M=53.12±22.36) in children. However, with respect to school functioning, the mean was observed to be low for all age groups at ages 5–7, 8–12, and 13–18, respectively (M=15.45±23.03; 46.50±25.98 and 52.142±4.32; p-value 0.01). The finding may suggest that the child’s perceived physical functioning affects the quality of life of pediatric cancer patients.

**Table 14.** Mean Scores of PedsQL™ 4.0 Generic Core Scale for Child and Parents proxy by age group (n=84)

| Domain | 5-7 yrs. |  | 8-12 yrs. |  | 13-18 yrs. |  | P-value |
| --- | --- | --- | --- | --- | --- | --- | --- |
|  | Mean score | SD | Mean score | SD | Mean score | SD |  |
| **Total mean score** | **57.32** | 12.34 | 70.00 | 15.32 | 73.33 | 10.31 | **<0.01** |
| Physical functioning | **53.12** | 22.36 | 71.59 | 19.09 | 71.14 | 19.99 | **<0.001** |
| Emotional functioning | 76.81 | 18.09 | 78.38 | 16.45 | 76.78 | 19.73 | 0.925 |
| Social functioning | 83.86 | 24.39 | 83.52 | 18.93 | 92.85 | 12.12 | 0.111 |
| School functioning | 15.45 | 23.03 | 46.50 | 25.98 | 52.14 | 24.32 | **<0.006** |

**Source:** Field Data (2022)

### Bonferroni alpha Post hoc test for health related quality of pediatric cancer patients for age

Multiple comparison tests in pediatric cancer patients showed a difference in significance between those aged 5-7 years and 8-12;13-18 years for physical and academic functioning, with the p-value being <0.01. While in the parents’ report, only the school function shows significant differences. However, in other domains, the observed mean difference was not statistically significant, as shown in Table 15. The results reflect that young pediatric cancer patients may face physical problems that can lead to developmental delays and limited school attendance and other problems.

**Table 15.** Bonferroni alpha Post hoc test for health related quality measured by (PedsQL™ 4.0) Generic Core Scale of pediatric cancer patients for age (n = 84)

| Variable | Comparison (year) | MD | S.E | 95% CI |  | P-value |
| --- | --- | --- | --- | --- | --- | --- |
|  |  |  |  | Lower | Upper |  |
| Physical functioning | 5 to 7- 8 to 12 | -18.47 | 5.38 | -31.62 | -5.33 | 0.003 |
|  | 5 to 7- 13 to 18 | -18.42 | 5.60 | -32.10 | -4.73 | 0.004 |
|  | 8 to 12-13 to 18 | -0.06 | 5.02 | -12.32 | 12.20 | 1.000 |
| Emotion functioning | 5 to 7-8 to 12 | -1.56 | 4.93 | -13.62 | 10.49 | 1.000 |
|  | 5 to 7- 13 to 18 | 0.03 | 5.14 | -12.52 | 12.59 | 1.000 |
|  | 8 to 12- 13 to 18 | -1.60 | 4.60 | -12.84 | 9.65 | 1.000 |
| Social functioning | 5 to 7-8 to 12 | 0.33 | 5.11 | -12.17 | 12.84 | 1.000 |
|  | 5 to 7- 13 to 18 | -8.99 | 5.32 | -22.01 | 4.02 | 0.285 |
|  | 8 to 12- 13 to 18 | 9.33 | 4.77 | -2.33 | 20.99 | 0.162 |
| School functioning | 5 to 7-8 to 12 | -31.05 | 6.76 | -47.57 | -14.53 | 0.000 |
|  | 5 to 7- 13 to 18 | -36.69 | 7.04 | -53.89 | -19.49 | 0.000 |
|  | 8 to 12- 13 to 18 | -36.69 | 6.30 | -9.76 | 21.05 | 1.000 |

Source: Field Data (2022)
MD-Mean Difference

### Mean scores of Health-related quality of life measured by (PedsQL 4.0) Generic core scale among pediatric cancer patients by hospitalization prior to 3 months

Pediatric cancer patients were hospitalized more than 3 times prior to 3 months, where the total mean scores were below 62.23±20.15. The overall trend physical functioning and school functioning decreased quality of life, as shown in Table 16.

**Table 16.** Mean scores of Health-related quality of life (PedsQL 4.0 Generic core scale) pediatric cancer patients by hospitalization before 3 months (n = 84)

| Domain | No Hospitalization |  | 1-3 times |  | ➤ 3 times |  | P-value |
| --- | --- | --- | --- | --- | --- | --- | --- |
|  | Mean score | SD | Mean score | SD | Mean score | SD |  |
| **Total mean score** | 79.71 | 30.47 | 72.33 | 21.16 | 62.23 | 20.15 | 0.192 |
| Physical functioning | 54.17 | 39.57 | 68.55 | 19.60 | 65.45 | 21.49 | 0.468 |
| Emotional functioning | 73.33 | 23.09 | 80.76 | 18.82 | 73.43 | 15.57 | 0.172 |
| Social functioning | 76.67 | 25.17 | 90.43 | 18.28 | 82.71 | 18.83 | 0.124 |
| School functioning | 48.33b | 34.03 | 49.57 | 27.93 | 27.31 | 24.71 | 0.002 |

Source: Field Data (2022)

Multiple comparison tests in pediatric cancer patients showed a difference in significance between those admitted 1-3 times and more than 3 times for school function 3 months before function by Bonferroni Alpha Post-Hoc Test with an adjusted p-value of <0.001 as shown in Table 17

**Table 17.**
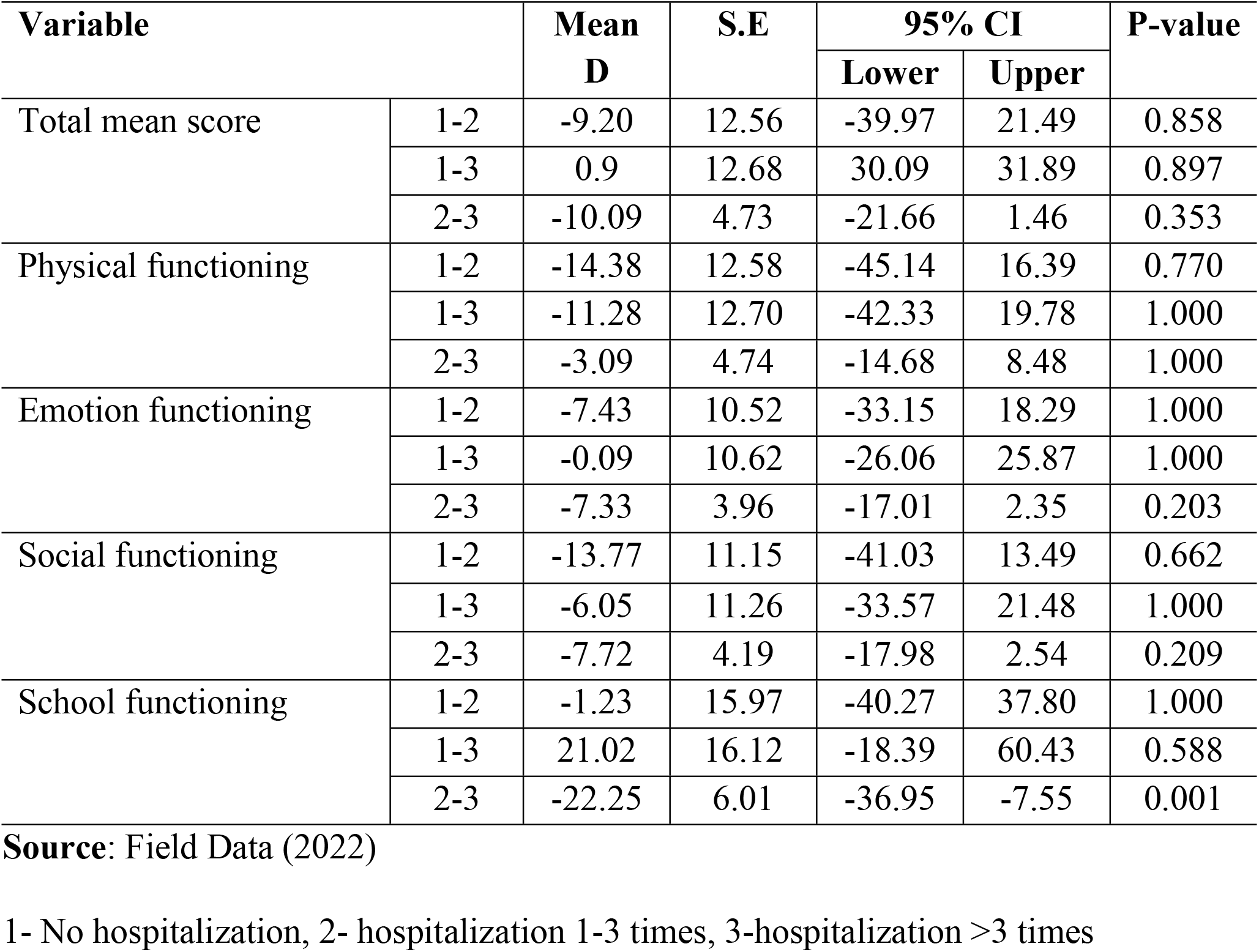
Bonferroni alpha Post hoc test of PedsQL™ 4.0 cancer generic scale with pediatric cancer patients for hospitalization prior in 3 months (n = 84)

| Variable |  | Mean<br>D | S.E | 95% CI |  | P-value |
| --- | --- | --- | --- | --- | --- | --- |
|  |  |  |  | Lower | Upper |  |
| Total mean score | 1-2 | -9.20 | 12.56 | -39.97 | 21.49 | 0.858 |
|  | 1-3 | 0.9 | 12.68 | 30.09 | 31.89 | 0.897 |
|  | 2-3 | -10.09 | 4.73 | -21.66 | 1.46 | 0.353 |
| Physical functioning | 1-2 | -14.38 | 12.58 | -45.14 | 16.39 | 0.770 |
|  | 1-3 | -11.28 | 12.70 | -42.33 | 19.78 | 1.000 |
|  | 2-3 | -3.09 | 4.74 | -14.68 | 8.48 | 1.000 |
| Emotion functioning | 1-2 | -7.43 | 10.52 | -33.15 | 18.29 | 1.000 |
|  | 1-3 | -0.09 | 10.62 | -26.06 | 25.87 | 1.000 |
|  | 2-3 | -7.33 | 3.96 | -17.01 | 2.35 | 0.203 |
| Social functioning | 1-2 | -13.77 | 11.15 | -41.03 | 13.49 | 0.662 |
|  | 1-3 | -6.05 | 11.26 | -33.57 | 21.48 | 1.000 |
|  | 2-3 | -7.72 | 4.19 | -17.98 | 2.54 | 0.209 |
| School functioning | 1-2 | -1.23 | 15.97 | -40.27 | 37.80 | 1.000 |
|  | 1-3 | 21.02 | 16.12 | -18.39 | 60.43 | 0.588 |
|  | 2-3 | -22.25 | 6.01 | -36.95 | -7.55 | 0.001 |

**Source:** Field Data (2022)
1- No hospitalization, 2- hospitalization 1-3 times, 3-hospitalization >3 times

### Health-Related Quality of Life and associated factors among Pediatric cancer patients

As shown in Table 18, regression analysis was performed to find the association between associated factors and HRQoL of pediatric cancer patients and their caregivers using the Pediatrics Quality of Life Inventory (PedsQLTM 4.0) Generic Core Scale. The results show a significant correlation between level of education, gender and age (=5.675; P-value <0.01: =3.685; P-value <0.01: =7.719; P-value <0.01). However, childhood deprivation (=-1.640; P-value<0.05), place of residence (=-3.131; P-value<0.05), work done in the last 7 days (=-6.476; P-value <0.01). hence, the finding implies that higher education, older age, and female gender increase the chance of an improvement in the quality of life. In addition, the chance of better health in pediatric cancer patients increases when childhood deprivation decreases, activities are reduced, and live in urban areas.

**Table 18:** Health related quality of life and associated factors measured by PedsQL™ 4.0 (n = 84)

| Variable | B | T | 95% CI |  | P-value |
| --- | --- | --- | --- | --- | --- |
|  |  |  | Lower | Upper |  |
| Child factors |  |  |  |  |  |
| Childhood deprivation | -1.640 | -2.211 | -3.116 | -0.165 | 0.030 |
| Place of residence | -3.131 | -0.219 | -5.797 | -0.464 | 0.022 |
| Age of a child | 7.719 | 5.105 | 4.710 | 10.727 | 0.001 |
| Sex of child | 3.685 | 1.451 | -1.368 | 8.737 | 0.001 |
| Level of education of the child | 5.675 | 4.395 | 3.103 | 8.246 | 0.000 |
| Work done in the past 7 days | -6.476 | -2.530 | -11.574 | -1.378 | 0.013 |
| Psychosocial counseling | -7.707 | -2.818 | -13.155 | -2.259 | 0.006 |
| Types of treatment received | -1.842 | -2.529 | -3.292 | -0.391 | 0.014 |
| Hospitalization in prior 3 months | -2.197 | -0.621 | -9.350 | 4.957 | 0.538 |
| Most activity done in the last 3 months | 1.611 | 1.247 | -1.001 | 4.223 | 0.220 |
| Caregiver factors |  |  |  |  |  |
| Level of education of the caregiver | -3.528 | -1.556 | -8.109 | 1.054 | 0.128 |
| Caregiver employment | 5.763 | 1.184 | -4.076 | 15.603 | 0.243 |
| Commence caring | -0.417 | -0.061 | -14.117 | 13.284 | 0.951 |

**Source:** Field Data (2022)

### Level of health-related quality of life among pediatric cancer patients measured by(PedsQL™ 3.0) Generic core scale

The results in Table 19 show that the mean total score (69.86±25.90) perceived by pediatric cancer patients corresponded to the cut-off point (69.7). The results suggest that pediatric cancer patients may have a poor quality of life.

**Table 19.** Total mean scores of Level of Health-Related Quality of Life (PedsQL 3.0) cancer module (n = 84)

| Domain | Mean | median | SD | Min | Max |
| --- | --- | --- | --- | --- | --- |
| **Total mean score** | 69.86 | 67.29 | 25.90 | 4.17 | 100.00 |
| Pain and hurt | 70.98 | 75.00 | 21.97 | 25 | 100.00 |
| Nausea | 76.49 | 80.00 | 18.54 | 0.00 | 100.00 |
| Procedural anxiety | 59.92 | 58.33 | 31.49 | 0.00 | 100.00 |
| Treatment anxiety | 84.92 | 100.00 | 23.33 | 0.00 | 100.00 |
| Worry | 77.48 | 75.00 | 22.73 | 0.00 | 100.00 |
| Cognitive problem | 62.20 | 77.50 | 37.29 | 0.00 | 100.00 |
| Perceived physical appearance | 83.63 | 100.00 | 24.42 | 8.33 | 100.00 |
| Communication | 48.81 | 50.00 | 27.48 | 0.00 | 100.00 |

Source: Field Data (2022)

### Mean scores of Health-related quality of life measured by PedsQL 3.0 cancer module perceived by pediatric cancer patients for sex

As shown in Table 20 independent t-test two tailed, the overall means are above the cut-off point and are not statistically significant. However, for procedural anxiety, cognitive problems, and communication, both male and female pediatric cancer patients had mean scores below their self-perceived limits. In addition, in the parent report, the mean score is lower in males (65.59±43.18) on cognitive function, although this was not significant. The finding implies that most male patients have a poor quality of life compared to female patients.

**Table 20.** Mean scores of Health-related quality of life (PedsQL 3.0 Generic core scale) perceived by pediatric cancer patients for sex (n = 84)

| Domain | Male |  | Female |  | P-value |
| --- | --- | --- | --- | --- | --- |
|  | Mean | SD | Mean | SD |  |
| **Total mean score** | 70.27 | 25.48 | 71.20 | 27.02 | 0.521 |
| Pain and hurt | 75.68 | 17.77 | 78.40 | 20.50 | 0.542 |
| Nausea | 71.39 | 22.27 | 70.00 | 21.65 | 0.814 |
| Procedural anxiety | 60.45 | 31.28 | 58.67 | 32.84 | 0.541 |
| Treatment anxiety | 83.89 | 24.84 | 87.33 | 19.56 | 0.901 |
| Worry | 77.68 | 20.56 | 77.00 | 27.67 | 0.485 |
| Cognitive problem | 60.33 | 37.53 | 66.60 | 37.13 | 0.329 |
| Perceived physical appearance | 85.88 | 23.53 | 78.33 | 26.13 | 0.197 |
| Communication | 46.89 | 26.03 | 53.33 | 30.71 | 0.364 |

Source: Field Data (2022)

### Mean scores of Health-related quality of life measured by PedsQL 3.0 cancer module perceived by pediatric cancer patients for age

As shown in Table 21 One-Way ANOVA Analysis of PedsQL Cancer Module 3.0. The finding shows that there is an association between health-related quality of life by age group where pediatric cancer patients ages 8-12 and 13-18 years have a high overall mean (74.31±12.32 and 75.15±9.74, respectively) that is statistically significant (p-value 0.01). The difference was seen in nausea (p-value 0.020), procedure anxiety (p-value 0.01), and cognitive problem (p-value 0.000) in the child report. Therefore, the results may suggest that aging may improve health-related quality of life in pediatric cancer.

**Table 21.** Mean scores of Health-related quality of life measured by PedsQL 3.0 cancer module for age (n=84)

| Domain | 5-7 yrs. |  | 8-12 yrs. |  | 13-18yrs. |  | P-value |
| --- | --- | --- | --- | --- | --- | --- | --- |
|  | Mean | SD | Mean | SD | Mean | SD |  |
| **Total mean score** | 58.90 | 11.37 | 74.31 | 12.32 | 75.15 | 9.74 | <0.01 |
| Pain and hurt | 64.77 | 23.98 | 73.89 | 22.05 | 72.32 | 19.94 | 0.296 |
| Nausea | 67.27 | 22.50 | 80.88 | 16.89 | 78.39 | 14.72 | <0.020 |
| Procedural anxiety | 71.96 | 33.09 | 87.99 | 16.30 | 91.36 | 17.49 | <0.01 |
| Treatment anxiety | 32.19 | 33.95 | 71.81 | 24.36 | 67.26 | 23.77 | <0.007 |
| Worry | 87.87 | 21.93 | 78.18 | 21.12 | 68.45 | 22.26 | <0.009 |
| Cognitive problem | 23.63 | 33.45 | 71.32 | 30.28 | 81.42 | 24.18 | <0.01 |
| Perceived physical appearance | 90.90 | 18.34 | 78.18 | 27.29 | 84.52 | 24.08 | 0.159 |
| Communication | 32.57 | 24.92 | 52.20 | 26.29 | 57.44 | 26.18 | <0.003 |

### Bonferroni alpha Post hoc test of PedsQL™ 3.0 cancer module scale of pediatric cancer patients for age

Table 22 Bonferroni alpha post hoc test for the PedsQL 3.0 cancer module in pediatric cancer patients aged 5-7, 8-12, and 13-18 years. The multiple comparison test revealed statistical differences between ages 5-7 and 8-13:5-7 and 13-18 for all domains except concern, with significance observed only between ages 5-7 and 13-18, with a p -score of <0.001 for procedural anxiety and cognitive problems with a p-value of <0.05 for nausea, worry, and communication. In parents/carers, post hoc tests show statistically significant anxiety, worry, cognitive, and communicative emotional functioning between children enrolled once or twice and children enrolled more than three times in relation to anxiety, worry, and communication.

**Table 22:**
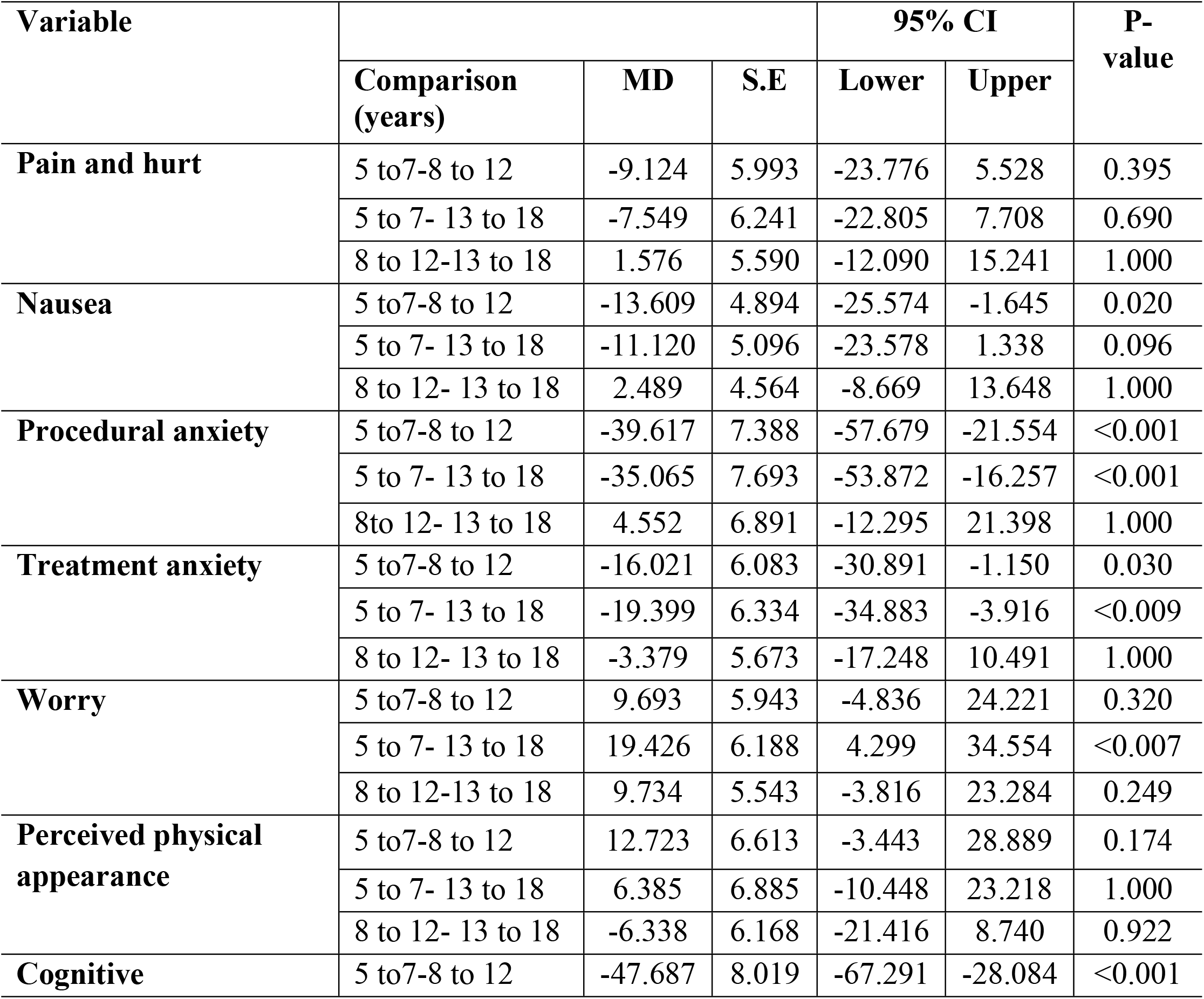
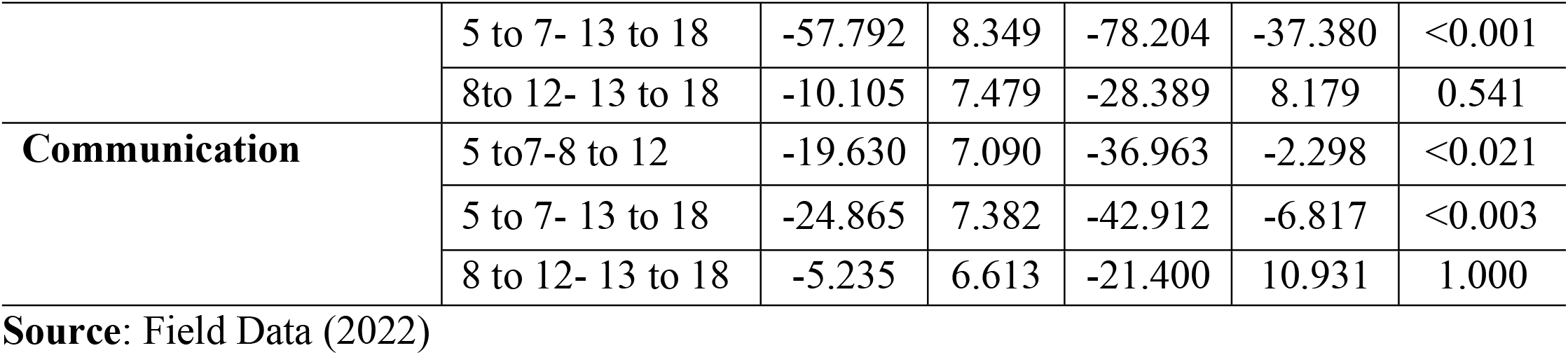
Bonferroni alpha Post hoc test of PedsQL™ 3.0 cancer module for age 5-7, 8-12, and 13-18 (n = 84)

### Associated factors and Health-Related Quality of Life among Pediatric cancer patients(PedsQL™ 3.0)

As shown in Table 23 a regression model to determine the effect of individual characteristics on the PedsQL 3.0 cancer module reports of children and parent proxy reports of quality of life. About PedsQL as perceived by pediatric cancer patients (child report). The finding indicates that the educational level of pediatric cancer patients is significantly associated with quality of life (=7.902 p<0.01), length of time since onset of disease (=2.165; p-value <0.01), and the number of siblings in the family (=2.488; p-value<0.05). The results suggest that a higher level of education, a longer survival time with cancer, and a large number of siblings increase the chance of a better quality of life. However, childhood deprivation included (=-2.175; p-value <0.01), complications related to cancer treatment (=-0.977; p-value <0.01), hospitalization in the past 3 months (=-747; p-value < 0.01).

**Table 23.** The health-related quality of life and associated factors (PedsQL™ 3.0 cancer module) (n = 84)

| Variable | B | t | 95% CI |  | P-Value |
| --- | --- | --- | --- | --- | --- |
|  |  |  | Lower | Upper |  |
| Child report |  |  |  |  |  |
| Childhood Deprivation | -2.175 | -3.122 | -3.561 | -0.789 | 0.002 |
| Place of residence | -4.573 | -2.658 | -8.051 | -1.096 | 0.011 |
| Age of the child | 1.796 | 0.770 | -2.920 | 6.512 | 0.446 |
| Sex of child | 3.506 | 1.150 | -2.658 | 9.670 | 0.257 |
| Level of education of the child | 7.902 | 7.302 | 5.747 | 10.056 | 0.000 |
| Work done in the past 7 days | -7.305 | -3.491 | -11.472 | -3.138 | 0.001 |
| Hospitalization in prior 3 months | -5.474 | -2.450 | -9.924 | -1.024 | 0.017 |
| Types of treatment the child received | -0.454 | -0.584 | -2.025 | 1.117 | 0.563 |
| Psychosocial counseling | -1.760 | -0.662 | -7.135 | 3.615 | 0.512 |
| Most activity is done in the last 3 months | 0.847 | 0.946 | -0.963 | 2.657 | 0.350 |
| **Parent/caregiver factors** |  |  |  |  |  |
| Level of education of the parent/caregiver | -0.589 | -0.375 | -3.764 | 2.585 | 0.709 |
| Employment | -0.823 | -0.244 | -7.640 | 5.995 | 0.809 |
| Commence caring | 15.677 | 3.338 | 6.185 | 25.169 | 0.002 |

**Source:** Field Data (2022)

In addition, results related to PedsQL perceived by parents/carers (parent report) show that childhood deprivation (=-1.589; p-value < 0.05), complications related to cancer treatment (=-1.159; p-value < 0.01)., hospitalization in the last 3 months (=-5.589; p-value <0.01). Therefore, the finding shows that reducing childhood deprivation, complications related to cancer, and the number of hospitalizations before 3 months increases the chance of an improvement in quality of life.

## Discussion

The present study found that there is association bweteen childhood deprivation and health related quality of life among pediatric cancer patients, while both level of childhood deprivation and health related quality were higher. However male pediatric cancer patients were many and suffering from Acute Lymphoblastic Leukemia.

The study results showed that acute lymphoblastic leukemia (ALL) and Hodgkin’s lymphoma were the most prominent leading cancers than other types of pediatric cancers. The most affected pediatric patients were males over 7 years of age compared to female patients. This is evident as many male pediatric cancer patients of school and adolescent age were compared with females in this study. The higher proportion of ALL and Hodgkin’s lymphoma observed in this study was similar to previous studies in South Africa which showed that various cancers were observed in which the majority of children diagnosed with acute lymphoblastic leukemia 12% and males were most affected [24]. However, the differences were found in latest study, the leading pediatric cancers were Wilms tumor 17.2%, retinoblastoma 16.8%, and acute lymphoblastic leukemia 15.9% [11]. There was a higher incidence of male subjects, according to international data reporting that males are more likely to have cancer than females, with a 6:5 ratio for males and females [18,39]. In addition, ALL was the most common cancer observed among all types of pediatric cancers [8,24].

In addition, many pediatric cancer patients were diagnosed in 2021 and early 2022 compared to previous years. This reflects that interventions such as childhood cancer awareness and community awareness programs were more widely implemented during this period than in previous years. In addition, priority was given to empowering healthcare providers through (short and long cancer training), in-service training, conferences, and seminars against pediatric cancer. Nervous was probably a time when there were advances in the diagnosis of pediatric cancer, resulting in a rising number of children being diagnosed and confirmed with cancer in Tanzania. However, only a few conducted studies reported that the mean time since diagnosis increased significantly after the advancement of pediatric cancer therapies such as multiple chemotherapies, surgery, radiotherapy, and psychosocial therapy [7]. Pediatric cancer patients were treated in more than 4 hospitals seeking treatment while the cancer symptoms were pre-diagnosis, with the majority of them, referred to MNH for diagnosis confirmation and treatment, resulting in late diagnosis and treatment as most of them took more than 2 weeks to arrive at the treatment center. This finding, also shown in previous studies, showed that about 74.5% of pediatric cancer patients live more than 4 hours to reach the cancer centers [11]. This is one of the factors contributing to being late in diagnosis, delaying cancer treatment can lead to early cancer complications, and early infant mortality since most pediatric cancer patients came from disadvantaged areas. [3,11], because of onset at a late stage of cancer.

Regarding the frequency of hospitalizations, most adolescent pediatric cancer patients were admitted 1 to 3 times prior to 3 months, which may be due to complications related to cancer itself and treatments. The result was consistent with a few studies conducted in which frequency of admission is related to concern [18]. Despite advances in cancer therapy that maximize survival time, pediatric cancer patients experience abdominal pain, fever, and palpitations during cancer treatment. As reported by many studies, most children with cancer face problems related to cancer and its therapy, including pain, nausea, vomiting, and physical incapacity leading to weakened immunity [28,29]. School/study events were the most activities performed by pediatric cancer patients in the last 3 months after play/excise and the few could not do any activities because they have a physical disability (handicap) due to cancer and they Delay in treatment and intervention.

In this study, the Multidimensional Poverty Index (MPI) was used to assess the level of childhood disadvantage in 84 of 91 pediatric cancer patients. The results of this study show that childhood deprivation was significantly higher in pediatric cancer patients as the proportion of deprived was larger than non-deprived, which is attributed to most of them missing their educational development while going to the hospital for treatment, frequency of admission, and informal employment of their parents and caregivers continued. The results were similar to previous demographic surveys: 50.2% of all children worldwide suffered from extreme poverty in terms of education, health, and nutrition, while 31% in sub-Saharan Africa were multidimensionally poor [15]. However, no significant difference was observed regarding the sex of the pediatric cancer patients, but the overall mean of the female patients was slightly higher compared to the male pediatric cancer patients, which may be due to the small number of female podiatry cancer patients participating in this study. However, about 12% of male children in Tanzania were disadvantaged in more than three essential needs, including early school dropping, health, and nutrition [15]

This study found that childhood cancer occurs in rural areas, has low levels of education, visits more than 4 hospitals while seeking treatment, and is significantly disadvantaged in single parent/carer living. However, the results showed a significant association between the standard of living and health. The difference was observed in place of residence between urban and rural and peri-urban and rural, while mean total scores were high for pediatric cancer patients living in rural areas compared to urban and peri-urban areas. Similar results of other studies from the 2016 NBS and UNICEF survey showed that about 81% of children living in rural areas are 3 and more disadvantaged, while few of them are in poverty (rural) areas compared to urban areas life [13]. In addition, children living in rural areas suffering from chronic diseases have been disadvantaged in more than basic needs compared to those living in urban [16]. Furthermore, few studies documented that most pediatric cancer patients living in non-deprived areas had more likely to have better health and improve their disease status to increase the survivership [40], compared to those coming from deprived areas or rural areas.

Time to start medication was significantly associated with childhood deprivation, as most pediatric cancer patients were delayed in arriving at cancer treatment centers. they took more than 2 weeks to start cancer medication after diagnosis of the types of cancer compared with previous finding access to cancer treatment centers and improving treatment modalities in national hospitals are important in delaying cancer treatment [3]. However, the findings suggested that there was no statistical difference either in gender or age among pediatric cancer patients as also reported in a previous study that there is no association between deprivation and gender or age group [41].

In this PedsQL study, the generic core module 4.0 was used to assess the health-related quality of life of 84 of 91 pediatric cancer patients aged 5 to 18 years whose parents/carers attended oncology clinics for treatment and other interventions.The results showed that the mean HRQoL scores perceived by pediatric cancer patients were lower in PedsQL inventory. Cancer is among the factors that have been perceived to contribute to the impairment of physical and school functioning in pediatric cancer patients. Male pediatric cancer patients are the most vulnerable group affected in both physical and school functioning, as their numbers were higher than female pediatric cancer patients in this study. Physical disability of pediatric cancer patients accompanied by school affection. Similar results were reported in previous studies, male pediatric cancer patients were the most prominent group discharged because of physical problems, seeking cancer treatment, and frequent admission to their school [18,24]

Furthermore, pediatric cancer patients aged 5-7 years were perceived as having physical and school functioning problems, while other groups were perceived only as having school functioning problems. HRQoL corresponding to age group implies that pediatric cancers are associated with better quality of life in older than younger pediatric cancer patients. In addition, most of them had not attended school because they did not feel well connected to cancer and its treatments and went to the doctor or hospital. However, participated in various activities in the last three months, including school/study, which is reliable with the current study not attending school for < months and continued in the last 30 days [24].

The similarity is also suggested by a study conducted in the US and Chile, in which pediatric cancer patients in Chile during the active phase of treatment and returning to school during their cancer treatment had a lower mean score. For all US school cancer treatment courses School functioning were the greatest impact on affecting the quality of life in pediatric cancer patients and the dissimilarity observed in the US, where there was a higher mean school functioning score throughout the treatment course [25]. The result suggests that there is a difference between countries’ scores on quality of life improvement and the impact of educational support. In addition, starting school and regular school attendance are believed to affect growth and development [10].

In this study, a linear regression analysis was performed to determine the impact of individual characteristics on HRQoL in pediatric cancer patients. Pediatric cancer patients, how were they disadvantaged, place of residence, participation in work/any activities in the last 7 days, their HRQoL were impaired resulting in poor quality of life (QoL). Poor health, the standard of living, and living in rural areas are the factors that contribute most to Poor QoL in pediatric cancer patients in this study. However, the results are consistent with recent studies suggesting that all children with cancer living in disadvantaged and rural areas are likely to impact poor QoL and experience a better quality of life if their socioeconomic status is improved [22,43].

However, the results obtained show that pediatric cancer patients who were female, school age, and adolescents (over 7 years) with higher education levels were perceived as having a better quality of life in both the child report and the parent proxy report. The results suggest that the age of pediatric cancer patients is more likely to affect the quality of life if it contributes to the normal developmental milestone even during the disease. However, younger age (5-7) years is associated with poor quality of life, which the child perceives in terms of physical functioning. This finding is the same as previous studies, which found that most pediatric patients diagnosed with cancer at a younger age had a poor observed quality of life [18,44]. This finding reinforces the importance of preventing the physical disability and dysfunction associated with cancer treatment and the disease itself.

In addition, the study was statistically significant in terms of perceived disease severity in pediatric cancer patients, who had lower overall mean. Procedural anxiety, cognitive problems, and communication problems were the critical problems perceived by pediatric cancer patients. Pediatric cancer patients rely on their parents/caregivers to provide their health information on their behalf. However, differences in three subscales, including nausea, procedural anxiety, and cognitive problems, were seen in the reports of children aged 8-12 and 13-18 compared to 5-7 years in this study. The observed outcome is no different from the current study, which found that younger-aged pediatric cancer patients were statistically associated with poor health-related quality of life on the fear of procedures subscale [18]. This finding underscores the importance of preventing the anxiety associated with medical intervention during cancer treatment. Procedural anxiety can be distressing pediatric cancer patients, making treatment difficult

Therefore, the results are likely contributed by other causes such as dissimilarity in developmental factors. In addition, the association was observed in the communication range, which was below the cutoff mean (69.7) in each age group. The same result was found in other studies that the communication range of pediatric cancer patients was considered difficult to interact with healthcare providers (Tembe, 2021). Additionally, most research on communication in childhood cancer has focused on parents/caregivers or healthcare providers rather than the patients themselves. Therefore, studies have suggested that understanding pediatric cancer patients require a healthcare provider to facilitate effective communication (patient-provider) which could improve quality of life and client satisfaction (Lin et al., 2020) through early intervention and reduce the impact of cancer therapies complication

The lower mean was found for pain and injury, nausea, and treatment anxiety perceived by pediatric cancer patients aged 5-7 years. In addition, a dissimilarity was found in the cognitive domain of the children’s report, where the mean score was lower at ages 5-7 than at ages 8-12 and 13-18, which is similar to other studies (Bishop et al., 2018; Tembe, 2021).. In addition, another analysis was performed (Bonferroni alpha post hoc test) to find out the difference between the age groups of the pediatric cancer patients, with the finding being consistent between the age groups (5-7 and 13-18; 5-7 and 13-18) significant was treatment anxiety. This may be related to a lack of patient orientation by healthcare providers and the developmental characteristics of children. In addition, cognitive problems have been observed between the ages of 5 and 7 years that being younger and being diagnosed with cancer can affect the quality of life, which is because most pediatric cancer patients do not attend school and spend most of their time in hospital spending.

In this study, a linear regression analysis was performed to determine the impact of individual characteristics on HRQoL in pediatric cancer patients. Pediatric cancer patients, how were they disadvantaged, place of residence, participation in work/any activities in the last 7 days, their HRQoL were impaired resulting in poor quality of life (QoL). Poor health, the standard of living, and living in rural areas are the factors that contribute most to Poor QoL in pediatric cancer patients in this study. However, the results are consistent with recent studies suggesting that all children with cancer living in disadvantaged and rural areas are likely to impact poor QoL and experience a better quality of life if their socioeconomic status is improved [22,44].

However, the results obtained show that pediatric cancer patients who were female, school age, and adolescents (over 7 years) with higher education levels were perceived as having a better quality of life. The results suggest that the age of pediatric cancer patients is more likely to affect the quality of life if it contributes to the normal developmental milestone even during the disease. However, younger age (5-7) years is associated with poor quality of life, which the child perceives in terms of physical functioning. This finding is the same as previous studies, which found that most pediatric patients diagnosed with cancer at a younger age had a poor observed quality of life [10,18]. This finding reinforces the importance of preventing the physical disability and dysfunction associated with cancer treatment and the disease itself. In addition, the regression model showed a significant association with quality of life (PedsQLTM 3.0 inventory) in both the child’s report and the parent’s proxy report.

The finding shows that the child’s age, hospitalization within the last 3 months, childhood deprivation, and school attendance were perceived as serious, and in the parent representative’s report the observed outcome was significant for hospitalization within the last 3 months and school attendance. The results may suggest that age, frequent hospitalizations, childhood deprivation, and irregular school attendance are important determinants of improved quality of life in pediatric cancer patients. However, most of the deprived pediatric cancer patients failed to attend school after frequent hospitalizations [44]. However, this study has discover that the childhood deprivation is higher which lead to lower quality of life among pediatric cancer patients related to delay treatment due to living in rural areas where there is limited of cancer centers and poor standard of living,

### Strength and limitation of the study

This study has discover that the childhood deprivation is higher which lead to lower quality of life among pediatric cancer patients related to delay treatment due to living in rural areas where there is limited of cancer centers and poor standard of living. The study did not rely on examining the clinical presentation factors associated with HRQoL. Also, the generality of this study may be limited by inclusion and exclusion criteria, particularly with regard to age, which did not involve pediatric cancer patients younger than 5 years of age and not able to understand and communicate verbally, and those who have been diagnosed with cancer and started or completed cancer treatment. Information bias can be observed after the Parent Proxy Report on Childhood Deprivation and Health-Related Quality of Life.

## Conclusion

The magnitude of acute lymphoblastic leukemia is higher among pediatric cancer patients. There was a significant link between pediatric cancers and childhood deprivation and thus, compromised quality of life among pediatric cancer patients. Innovative pediatric cancer care policies, guidelines, and or strategies may need to be advocated to address the problem accordingly.

## Suggestions

Future research to maximize the size of the study by recruiting a large sample is needed to investigate routine clinical factors that may affect improving QOL outcomes using sensitivity measures during the treatment of pediatric cancer patients that provide important information for early information of healthcare providers about treatment decisions represent intervention.

## Data Availability

All relevant data are within the manuscript and Supporting Information files

## Supporting information

Dataset available, shared upon request

## Acknowledgments

The authors express gratitude to the University of Dodoma for providing ethical clearance to conduct this study. We also thank the Muhimbili National Hospital administration for allowing us to conduct the study, also we thank the pediatric cancer patients and their parents/caregivers for volunteering to participate in this study fully till the end.

## Author Contributions

Conceptualization: Mwanaheri Chubi.

Data curation: Mwanaheri Chubi

Formal analysis: Mwanaheri Chubi.

Methodology: Mwanaheri Chubi.

Supervision: Stephen Kibusi, Lulu Chirande and Shakiru Jumanne.

Varidation : Mwanaheri Chubi

Writing: original draft: Mwanaheri Chubi.

Writing: review & editing: Stephen Kibusi, Lulu Chirande and Shakiru Jumanne.

Funding: This research received no external funding

